# Extended-Spectrum Beta-Lactamase *Klebsiella pneumoniae* on a Malawian neonatal unit is amplified by neonates and transmitted by maternal hands, cots and ward surfaces

**DOI:** 10.1101/2025.08.13.25333346

**Authors:** Oliver Pearse, Rebecca Lester, Allan Zuza, Helen Mangochi, Patricia Siyabu, Edith Tewesa, Thomas Edwards, Nicholas R. Thomson, Kondwani Kawaza, Patrick Musicha, Jen Cornick, Eva Heinz, Nicholas Feasey, Chris Jewell

## Abstract

**Background:** *Klebsiella pneumoniae* (*Kpn*) is an important cause of neonatal sepsis in sub-Saharan Africa. *Kpn* are intrinsically resistant to penicillins and frequently resistant to gentamicin and 3^rd^-generation cephalosporins, key agents in the treatment of neonatal sepsis. Such infections can be prevented by effective infection prevention and control (IPC), but the most effective IPC packages for resource-limited healthcare settings are incompletely defined.

**Methods:** We recruited a cohort of 94 neonates admitted to a Malawian neonatal unit, alongside their mothers, and utilised single colony whole genome sequencing and post-enrichment metagenomics to determine the most important transmission routes of Extended-Spectrum Beta-Lactamase producing (ESBL) *Kpn*. These data were analysed at the level of ST and pairwise SNP distance and combined with statistical models to infer transmission.

**Findings:** ESBL-*Kpn* rapidly colonised neonates; female sex, receipt of oxygen, caesarean delivery and antibiotic use increased colonisation risk. STs causing invasive infection and stool colonisation were temporally related to those found in the ward. There was greatest circulation of ESBL-*Kpn* between compartments that are in contact with neonatal stool (neonate stool, mothers’ hands, cots, and the swaddling cloths). Utilising epidemiological and SNP data, *Kpn* appeared to be transmitted to neonates primarily from cots, ward surfaces (sinks and oxygen delivery equipment) and from other neonates. Network analysis implicated cots, antibiotics and oxygen delivery in ESBL-*Kpn* transmission.

**Interpretation:** An unsafe hospital environment is strongly implicated in neonatal invasive infection and stool colonisation with ESBL *Kpn*. IPC interventions should focus on containing neonatal stool, hand hygiene, cot decontamination, single use oxygen delivery and surface cleaning, particularly sinks.

**Funding:** This work was funded by the Antimicrobial Resistance Cross-Council Initiative through grants from the Medical Research Council, a Council of UK Research and Innovation and the National Institute for Health Research (MR/R015074/1 & MR/S004793/1); and the Bill and Melinda Gates Foundation (INV-005692). Malawi-Liverpool-Wellcome Research Programme (MLW) is core-fund by Wellcome (206545/Z/17/Z).

**Research in context:** *Evidence before this study:* Neonatal sepsis is a leading cause of neonatal mortality, particularly in sub-Saharan Africa. *Klebsiella pneumoniae* (*Kpn*) has emerged as an important cause of neonatal sepsis and is difficult to treat due to increasing antimicrobial resistance. Extended-spectrum beta-lactamases are enzymes that confer resistance to ceftriaxone, and for neonates in many sub-Saharan African neonatal units this resistance determinant renders their infections untreatable. While infection prevention and control (IPC) measures are important to limit transmission, the most important IPC measures in this context remain unclear.

*Added value of this study:* This study shows that colonisation with ESBL *Kpn* in the neonatal unit is common. Risk factors for colonisation include oxygen delivery, antibiotic therapy, and female gender. Isolates from invasive disease were genomically like those that caused stool colonisation and that were colonising the general ward environment. There was circulation of bacteria between the neonatal gastrointestinal tract, cots, mothers’ hands, and swaddling cloths. Transmission appeared to be from cots, ward surfaces, particularly sinks and oxygen equipment, other neonates, and the mothers’ hands.

*Implications of all available evidence:* IPC should be prioritized to reduce colonisation and subsequent infection with ESBL *Kpn*, and other enteric pathogens regardless of AMR profile. Suggested IPC interventions in this context should focus on reducing bacterial load in the ward environment, with neonatal stool management, maternal hand hygiene, single-use oxygen delivery equipment, replacing sinks with hand sanitizer and cot decontamination.

## Introduction

Neonatal infection (primarily sepsis and meningitis) is the third greatest cause of neonatal death globally^1^, with *K. pneumoniae* (*Kpn*) a leading cause of healthcare-associated neonatal sepsis^2^. In sub-Saharan Africa (sSA)^3,4^, the area which carries the highest burden of neonatal sepsis, *Kpn* infection is particularly problematic given the high prevalence of antimicrobial resistant (AMR) lineages resistant to first- and second-line agents for neonatal sepsis (penicillins, gentamicin and ceftriaxone). Key AMR determinants are Extended-Spectrum Beta-Lactamases (ESBLs), which inactivate 3^rd^-generation cephalosporins (3GC) like ceftriaxone. Ceftriaxone is often second-line therapy for neonatal infection and, in resource-limited healthcare settings, may be last-line, leaving these infections *de facto* untreatable. Other Gram-negative bacteria commonly causing neonatal sepsis that are also 3GC-resistant include *Escherichia coli, Acinetobacter baumannii* and *Enterobacter cloacae*.

Given high levels of treatment failure in neonatal sepsis^5^, the consequent mortality and the fact that for pathogens such as *Kpn* a large proportion of infections are healthcare associated (HCAI)^6^, attention is increasingly focusing on improving Infection Prevention and Control (IPC) programmes. There is, however, limited data on optimal IPC strategy to reduce neonatal infection in low-income settings^7^. In some settings, mothers provide much of the neonatal care, neonates may share cots due to limited resources, and components of medical equipment—such as oxygen tubing—that are difficult to sterilize may be reused. Additionally, variations in swaddling and nappy use, along with inconsistent access to running water, further complicate infection prevention. Consequently, IPC practices effective in high-income neonatal units may be impractical or ineffective in sub-Saharan Africa.

Since gut mucosal colonisation with AMR organisms such as ESBL-*Kpn* is a key risk factor for invasive disease^8^, studies of enteric *Kpn* transmission have considerable potential to inform IPC practice. Here, we investigate inpatient neonatal gut mucosal colonisation with ESBL *Kpn* on a tertiary referral Malawian neonatal unit (NNU) to determine the most important transmission routes of ESBL-*Kpn* to inpatient neonates, with a focus on guiding IPC practice.

## Methods

### Study site

Queen Elizabeth Central Hospital (QECH) is a government-run tertiary hospital for Blantyre and southern Malawi, providing free healthcare. Its maternity services are comprehensive, with 30% of ~14,000 births delivered by caesarean section^9^ and a stillbirth rate of 3.4%^10^. Chatinkha nursery (Supplementary Figure 1), the NNU, admits ~5,000 neonates born annually at QECH, 80% of which are born at QECH. In 2013, neonatal mortality on Chatinkha was 17%^11^. The nursery offers oxygen therapy, continuous positive airway pressure, intravenous medication, and phototherapy; surgical cases are referred to the on-site surgical hospital, which provides intensive care. Neonates are reviewed daily by doctors or clinical officers, with 3-6 neonatal nurses on duty per shift.

### Study details

This prospective cohort study was conducted on the Chatinkha nursery. Infants were recruited within 48 hours of admission and followed up to day seven or discharge. Infants were eligible if admitted to the nursery, expected to stay >24 hours, and had a consenting guardian aged ≥18 years. Data from medical notes and caregivers were collected using Open Data Kit (ODK)^12^.

Samples were either mother & baby dyad associated or from the broader healthcare environment. Neonate-associated samples (stool/rectal swabs, maternal stool/rectal swabs, maternal hand swabs, cot swabs, and swaddling cloth samples) were collected at admission, day three, and day seven or discharge. In addition, 20 pre-determined environmental samples from ward surfaces and pooled staff hand swabs were sampled weekly (Supplementary Figure 1 and Supplementary methods). Neonate and cot movements were tracked using QR codes placed on cots, wristbands, and walls, scanned twice daily to build a dynamic map of neonate and cot locations over time.

Study samples were processed in the laboratory using ESBL selective media to isolate ESBL *Kpn* and Gram-negative selective media for plate sweeps to capture overall *Kpn* diversity. Both isolates (single colony WGS) and plate sweeps (post-enrichment metagenomics^13, 14^) underwent DNA extraction and Illumina next generation sequencing and were analysed bio-informatically (Supplementary methods and published elsewhere^15^). Clinical isolates were identified (Supplementary methods and published elsewhere^15,16^) and underwent the same DNA extraction and Illumina sequencing protocols as study isolates.

### Statistical analysis

The methods are summarised below with a full description of the statistical analysis in the Supplementary methods.

### State transition model

Colonisation status of neonates and their mothers was modelled using a chain-binomial discrete time Markov process^17^ using a Metropolis-within-Gibbs MCMC algorithm using the gemlib library^18^ (code on GitLab^19^). Briefly, we considered neonates and their mothers to exist in one of two mutually exclusive states, either uncolonised (*U*) or colonised (*C*). We denote 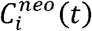 and 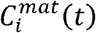 as a neonate and mother *i*’s colonisation status respectively at time *t* (in days), taking the value one if colonised and zero otherwise. We used MCMC to model the rate of transition from *U* to *C*, the effect of mothers on neonatal transition and the effect of covariates on this rate.

### Source attribution model

The source attribution model employed case counts by Multi-locus Sequence type (ST) and sample type in a Bayesian implementation of a modified Hald model^20^ (Supplementary methods). Neonatal stool colonisation episodes with each ST are modeled as a Poisson variable, influenced by the ST’s abundance in various compartments, the effect of each source, and a “strain” effect for each ST.

### Transmission analysis

Transmission analysis was conducted using WGS data by defining a SNP threshold, below which isolates were considered likely clonal (seven SNPs; Supplementary methods). Ward compartments were defined, and the density of circulation between them was assessed. Using the SNP threshold and sample timing, putative transmission events for each neonatal colonisation episode were identified. Sensitivity analysis was performed using SNP thresholds of zero, three and 20.

### Exponential Random Graph Models (ERGMs)

ERGMs were implemented using the R packages detailed in the Supplementary methods. Two networks were defined: one based on neonates with a pairwise SNP distance between their *Kpn* isolates ≤ the threshold, and another linking neonates by contemporaneous or sequential cot occupancy. The presence of an edge in the first network was regressed on the presence of an edge in the second network, time difference between samples, overall network density, and nodal covariates.

### Ethical approvals

Ethical approval for the study was granted in Malawi by Kamuzu University of Health Sciences REC, College of Medicine Research Ethics Committee (COMREC P.10/18/2499) and LSTM REC (19-018). LSTM acted as the study sponsor.

## Results

From July 2019 to April 2020, we recruited 94 neonates (Supplementary Figure 2); 12/91 (13.2%) with outcome data available died, 64/91 (70.3%) survived to discharge and 15/91 (16.5%) were in-patients at last follow-up. Median gestational age was 37 weeks, with 38/94 (40%) < 37 weeks of gestation (Table 1). Average birthweight was 2535g with 38/94 (40%) cases low birthweight (LBW). Median age at recruitment was one day. Mothers received antenatal care (primarily in governmental clinics) in 86/94 (91%) cases and 15/94 (16%) were HIV-infected. Mode of delivery was caesarean section for 18/94 (19%). Place of delivery was QECH for 65/94 (69%) neonates, with the rest born at healthcare centres, other hospitals or on the way to hospital. 30/94 (32%) mothers were exposed to antibiotics, either prenatally (18/94 [19%]) or intrapartum (24/94 [24%]; Supplementary Table 1). Most neonates 69/94 (73%) received antibiotics, most frequently benzylpenicillin/gentamicin 66/94 (70%), with fewer receiving ceftriaxone, the second-line therapy 6/94 (6%).

**Table 1.**
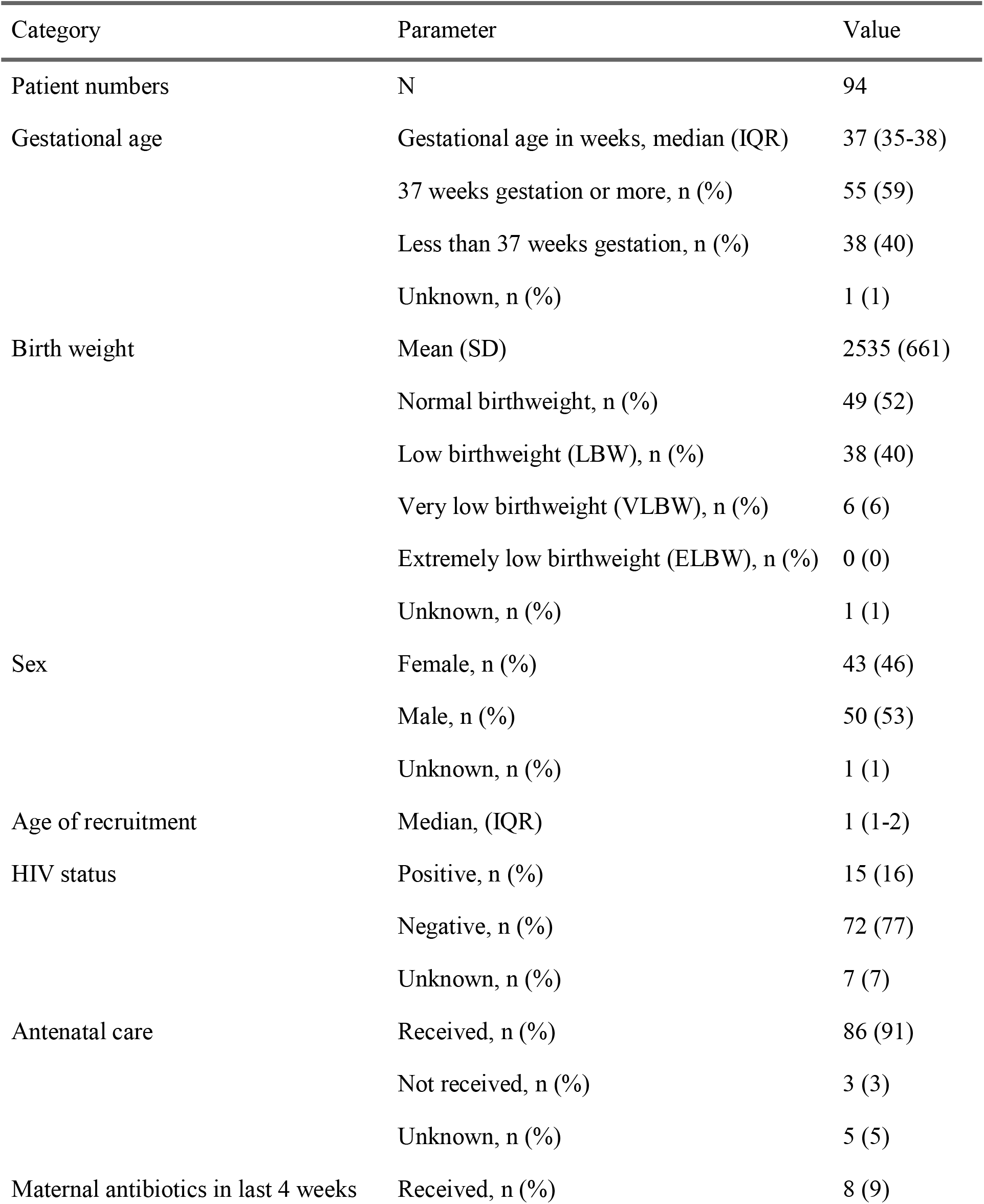
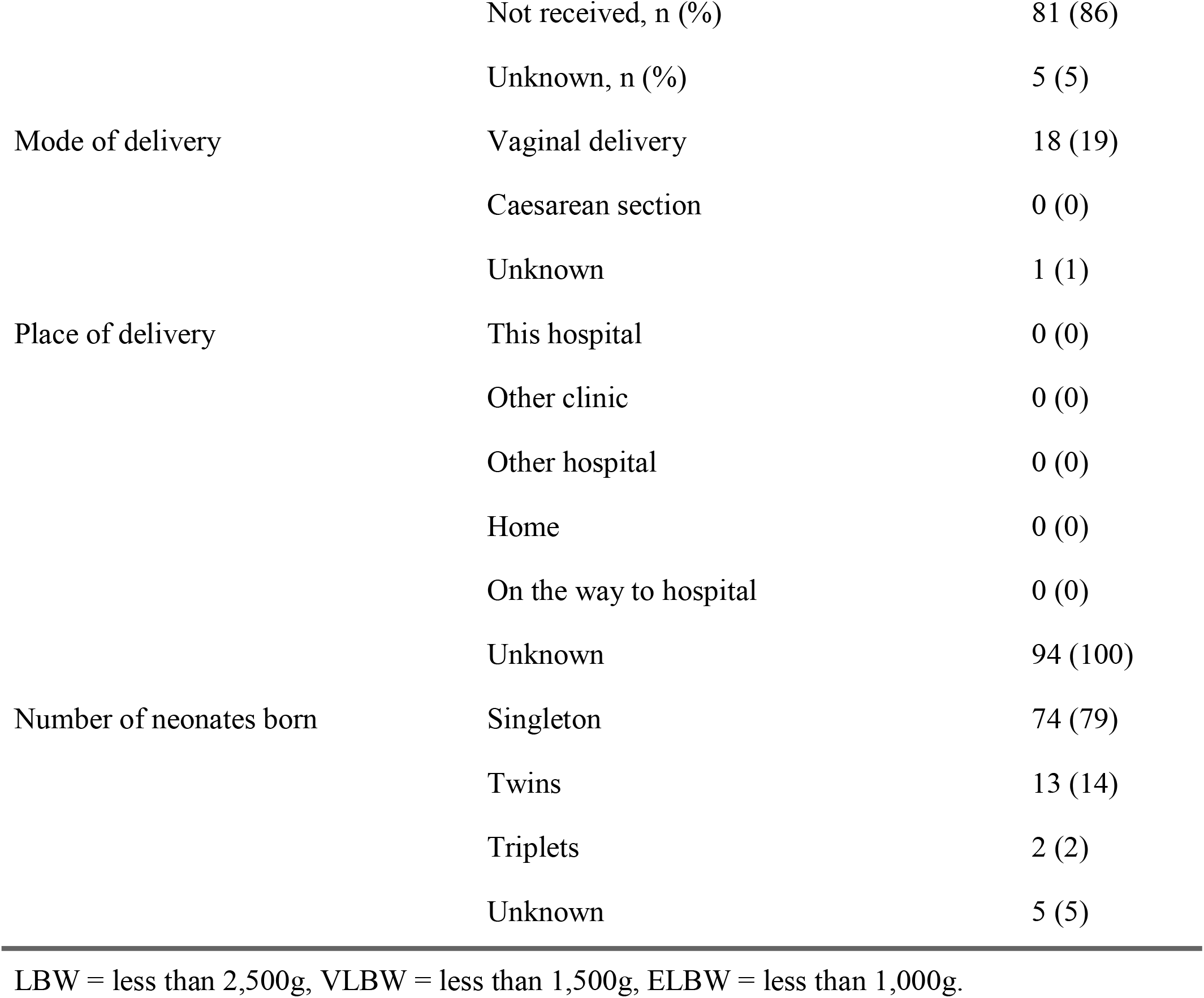
Clinical characteristics of the neonates and mothers enrolled in the study.

During admission, 64/90 (71%) neonates were colonised by an ESBL organism at least once (Figure 1A). Median time to ESBL colonisation was three days [95% CI 2 - 4], and 94% [95% CI 81% - 98%] were colonised by seven days. Neonatal colonisation by ESBL *Kpn* was more rapid (median five days [95% CI 4 - 6]) than any other ESBL organism with 69% [95% CI 53% - 80%] colonised at seven days (Figure 1B). In contrast, mothers were more likely to be colonised by ESBL *E. coli* (22% greater proportion of maternal stool positive for *E. coli* compared to *Kpn* [95% CI 11% - 32%; *p* < 0.0001]). Maternal hand swabs (85% [95% CI 80% - 90%]) and surface swabs (84% [95% CI 81% - 87%]) were most likely to test positive for ESBL *A. baumannii* (Supplementary Figure 3).

**Figure 1.**
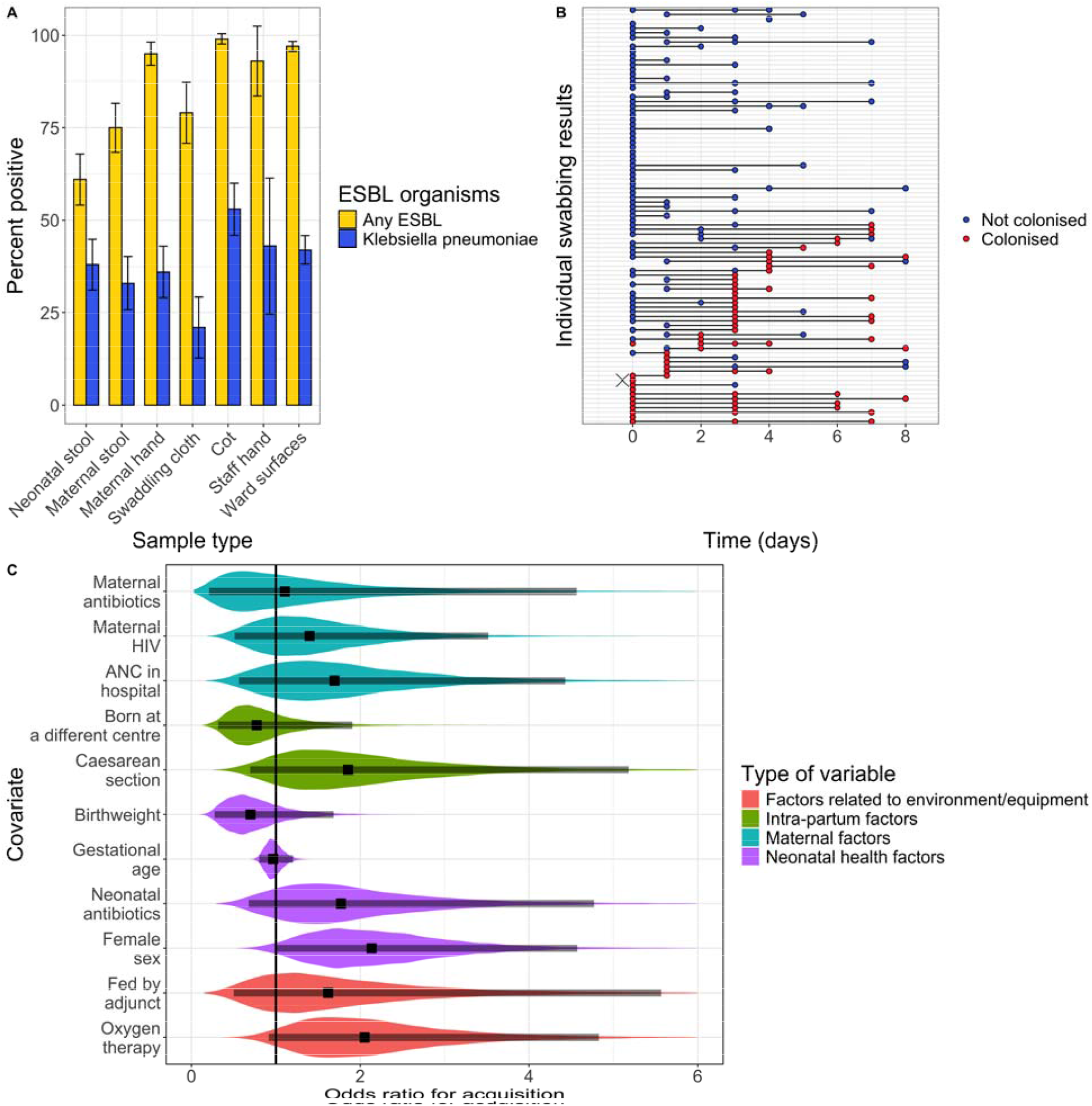
A) The percent of all samples that were positive for any ESBL organism (yellow) or ESBL-Kpn (blue) by sample type. B) The individual swabbing results of all neonates recruited into the study. Each row represents a neonate, each point represents a swabbing event, with red indicating the neonate as colonised with ESBL-Kpn and blue indicating they are not colonised with ESBL-Kpn. The cross represents the neonate which had invasive Kpn sepsis. C) A violin plot showing the distribution of the posterior of the neonatal and maternal covariates, arranged by group. The violins represent the overall posterior distribution, the points represent the median value and the lines represent the 95% credible intervals (CrI).

In our state transition risk factor model the baseline rate for neonatal ESBL-*Kpn* colonisation was 0.13 [95% Credible Interval (CrI) 0.07 - 0.20] per day, and 0.10 [95% CrI 0.06 - 0.16] for mothers. The increase in the rate of neonatal colonisation if their mother was colonised (Φ) was 2.18 [95% CrI 0.54 – 5.40]. Female sex (HR 2.14 [95% CrI 1.01 – 4.57]), receipt of oxygen therapy (HR 2.05 [95% CrI 0.92 – 4.83]), caesarean delivery (HR 1.86 [95% CrI 0.70 – 5.18]) and neonatal antibiotics (HR 1.77 [95% CrI 0.68 – 4.77]) increased the risk of becoming colonised by ESBL *Kpn* (Figure 1C), though only the credible intervals for female sex did not cross one. On day three of life (sampling tended to be on day 4) the predicted colonisation was 14% greater than observed, implying that interval censoring by sampling infrequently underestimates speed of colonisation.

We included all genomic data from single colony WGS and pseudo-assemblies (Supplementary methods) identified as *K. pneumoniae sensu lato* for genomic analysis based on ST, and *K. pneumoniae* subsp. *pneumoniae* for genomic analysis based on SNPs (Supplementary methods).

We examined the interplay between *Kpn* causing invasive infection and stool colonisation and *Kpn* found in the ward environment. Twenty-six *Kpn* were isolated by routine diagnostic microbiological culture from blood or CSF of neonates during the study; of which 17 ceftriaxone resistant isolates were analysed further (Supplementary methods). There was positive correlation between the number of invasive isolates and stool colonisation isolates of each ST identified during our study period (*ρ* = 0.47, *p* < 0.00001), indicating that the ward level population of stool colonisation isolates is representative of the invasive isolates. 10/11 (91%) of *Kpn* STs (corresponding to 16 isolates) causing invasive disease were found in the ward, suggesting *Kpn* in the ward environment was contributing to invasive disease (Figure 2A). One invasive ST584 isolate was from a study participant; this ST was also found in the neonate’s stool, cot and swaddling cloth and the mother’s stool and hands.

**Figure 2.**
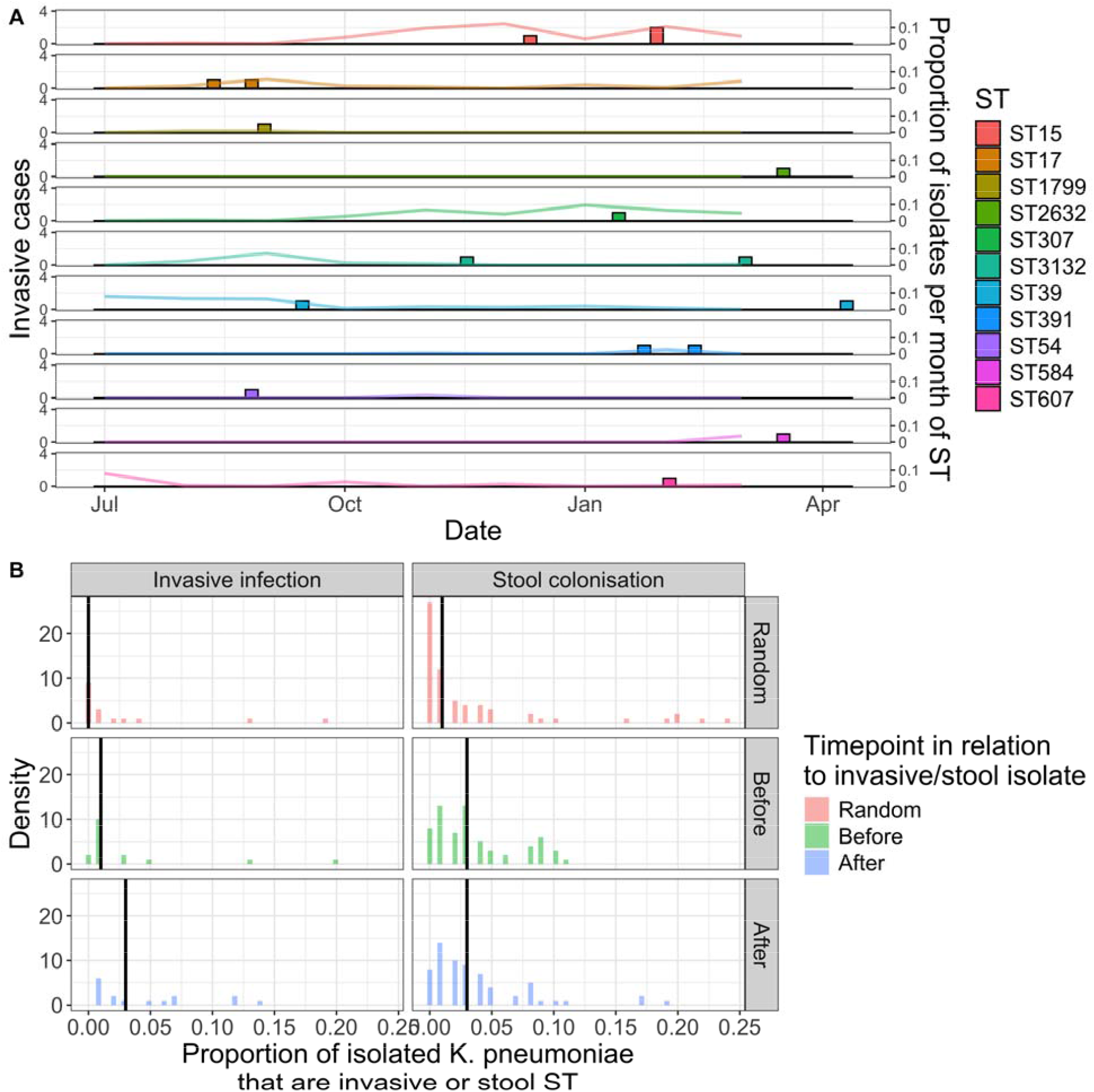
A) A timeline showing the occurrence of invasive isolates of each ST on the ward (bars), alongside the proportion of all samples on the ward that were positive for that ST (lines). B) A frequency chart showing the proportion of study samples of a specific ST isolated in the 28 days before (green), after (blue) and at a random time in relation to it (red). STs shown are either those that were invasive isolates or stool colonisation in the neonate.

For all *Kpn* isolates causing invasive disease and stool colonisation in neonates, we examined three timepoints per isolate; 28 days before and 28 days after isolation, and a randomly selected 28-day window during the study period. For each timepoint, sample-positivity for any given ST derived from an invasive or colonisation sample was calculated as a proportion and compared between groups. Median proportion of the respective ST in a randomly selected 28-day period was lower (0) than in the 28-days before an invasive isolate (0.01 [*p* = 0.05]) or 28 days immediately after (0.03 [*p* = 0.004]; Figure 2B). For stool colonisation, the median proportion of the respective ST in a randomly selected 28-day period was lower (0.008) than in the 28-days before stool colonisation (0.03 [*p* < 0.002]) or in the 28-days after stool colonisation (0.03 [*p* < 0.003]; Figure 2B) suggesting spillover from patients into the ward environment. This relationship was also observed for metagenomic stool samples when analysing the mSWEEP post-enrichment metagenomic data, but not for the invasive isolates (Supplementary Table 2).

We utilized a source attribution model based on ST and a SNP based analysis to determine circulation and transmission of *Kpn around* the ward. Based on ST frequency in single colony WGS isolates in our source attribution model, the relative contributions to neonatal stool colonisation were greatest for the cots 0.67 (95% CrI 0 - 0.95) and the mother’s hands 0.17 (95% CrI 0 – 0.60; Figure 3A), however when the mSWEEP data was used the contribution estimated due to cots increased to 0.96 (95% CrI 0.45 – 0.99) and the contribution estimated due to mother’s hands decreased to 0 (95% CrI 0 – 0.30; Supplementary Table 3).

**Figure 3.**
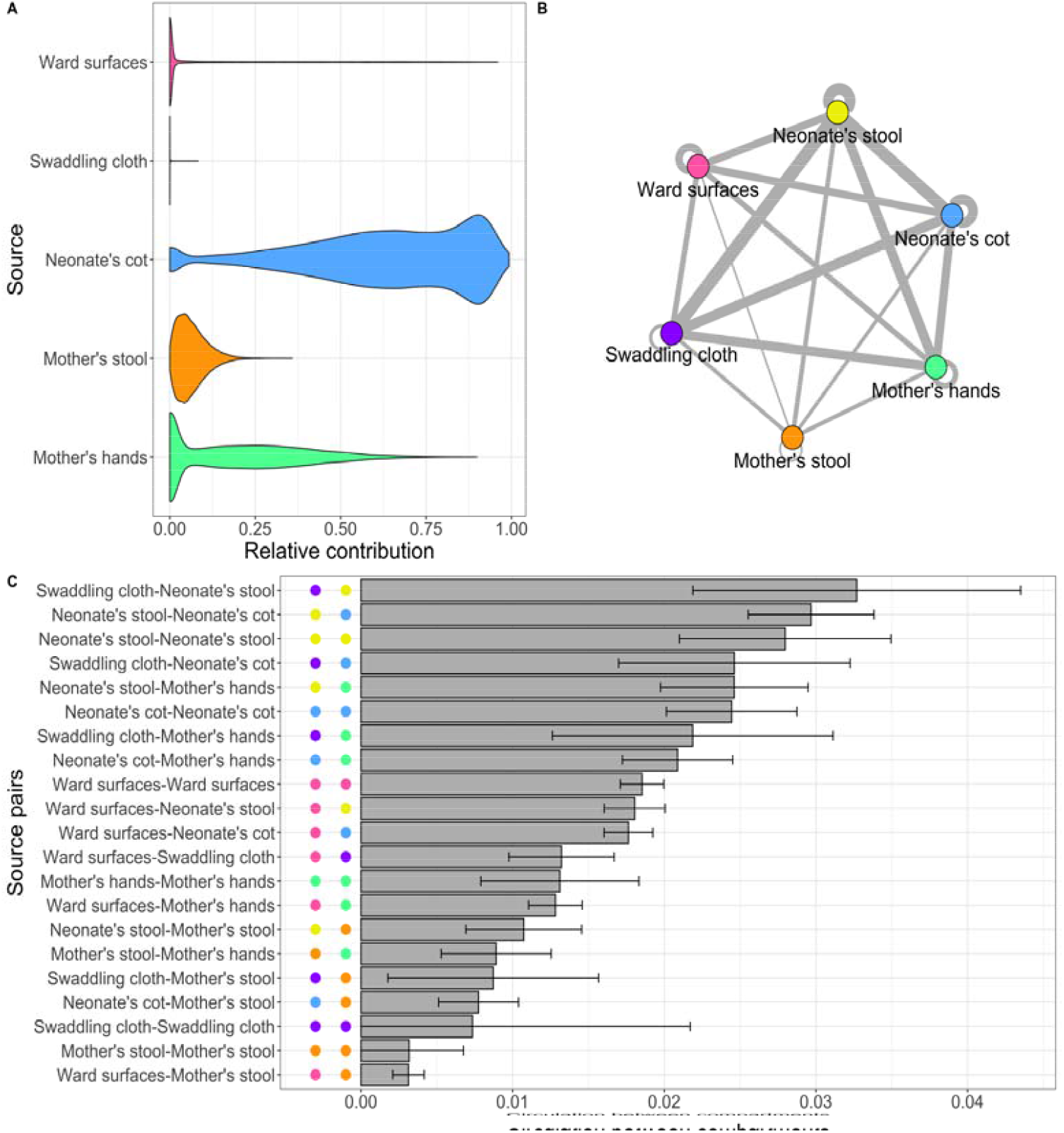
A) A violin plot showing the relative contributions of different sample types to neonatal colonisation based on relative frequencies of STs in each sample type. B) A network graph representation of Figure 3C showing circulation between different compartments. Each node represents a different compartment, with the edges representing movement between them. The thickness of the edges represented movement between two compartments (in either direction). C) Circulation between the different ward compartments, as defined by a pairwise SNP distance between isolates of seven or less. Circulation is determined as the number of isolates between two compartments with a pairwise SNP distance below the threshold divided by the total number of potential connections between the two compartments. The colour of the points represents the two sources in the pair for that bar.

We examined mixing between different ward compartments (Figure 3B & 3C), using a threshold of seven SNPs to identify circulation of *Kpn* around the ward (Supplementary methods & Supplementary Figure 4). To determine this the number of transmission links were divided by the total number of potential connections between sources. The greatest movement was between and within swaddling cloths, neonates’ stool, and neonates’ cot compartments (density of circulation from 0.021 – 0.027). There was also high movement between mothers’ hands and neonates’ stool (density of circulation 0.022), and within the ward surface compartment (density of circulation 0.016). The least movement was between mothers’ stool and the other compartments (0.0029 – 0.0089). Analysis of metagenomic data (mGEMS) and sensitivity analysis using a varying SNP threshold showed similar results, though suggested more circulation between the mother’s stool compartments and other compartments (Supplementary Figure 5 & 6).

To investigate the role of different sources of transmission to neonates, colonisation episodes (CEs) were examined using single colony WGS. Most but not all neonates were colonised by just one ST; each colonisation with a single ST was defined as a separate CE and analyzed separately, resulting in 52 CEs for 41 neonates. We defined putative transmission events for a neonatal CE by comparing the CE isolate with isolates on the ward of all sample types 1-14 days before the CE commenced and identifying isolates with a pairwise SNP distance ≤ seven. Some STs were found on the ward throughout the study (ST15, ST2793), but with a few exceptions’ lineages tended to occur in short-lived clusters that re-emerged rather than being present constantly, making a 1-14 day time window appropriate for analysis (Supplementary Figure 7). We found no putative transmission events for 28 CEs, but for the 24 CEs where putative transmission events could be identified, median putative transmission events/CE was two and one CE had 15 (Figure 4A & 4C). The most frequent putative sources of transmission were ward surfaces (33/95 [34.7%]), followed by cots (29/95 [30.5%]), other neonates (15/95 [15.8%]) and mother’s hands (10/95 [10.5%]). Utilising post-enrichment metagenomic assemblies (mGEMS), ward surfaces were even more strongly implied as a likely source, representing 49/74 (66.2%) of all putative transmission events. Within the ward surface swabs, sinks emerged as a putative source most frequently, with 15/33 (45%) and 16/49 (33%) transmission events based on single colony WGS and post-enrichment metagenomics, respectively, whilst 9/49 (18%) transmission events were from oxygen delivery or CPAP equipment using post-enrichment metagenomics. Sensitivity analysis with SNP distances between zero and 20 revealed similar effects, though with far fewer transmission events at zero SNPs (Supplementary Table 4).

**Figure 4.**
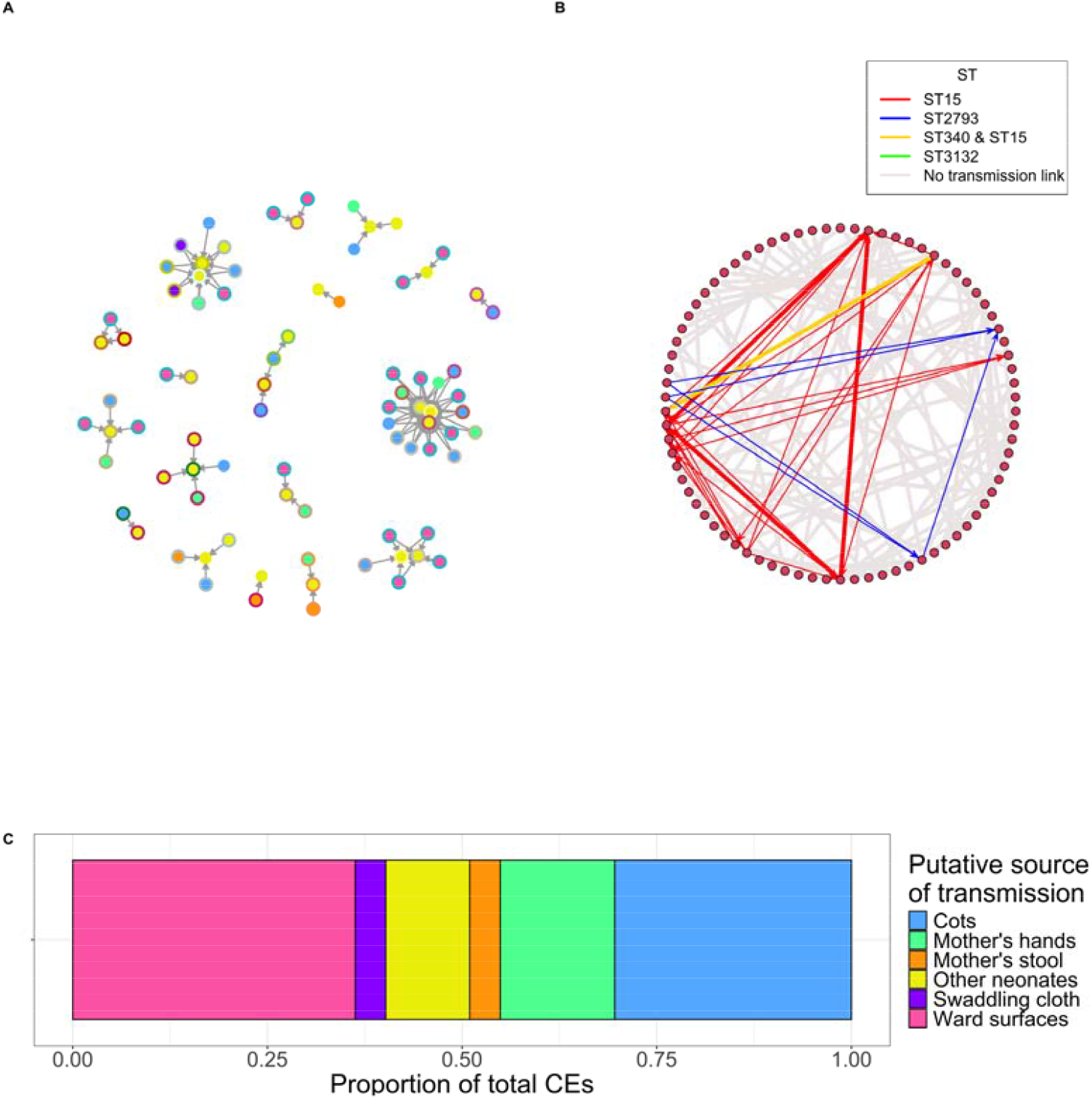
A) A network graph showing putative transmission events, the internal node colour represents the sample source, and the external border colour represents which dyad the isolate belongs to. Grey external borders represent isolates coming from general ward surfaces. B) A network graph showing the cot network superimposed on the transmission network. Nodes represent individual neonates. Thick arrows represent neonates that occupied a cot contemporaneously or sequentially, with the arrow pointing to the neonate that occupied the cot second. Coloured thin arrows represent neonates that are connected by potential TEs, but not connected by cot. Coloured thick arrows represent neonates that are connected by potential TEs and also by cot. The colour of the arrows represents which ST the neonates were colonised with. Grey arrows represent neonates connected by cot that did not have a potential TE. C) A Bar chart representing the proportion of transmission events ascribed to different sources.

To formally test whether transmission links were related to neonates occupying the same cots, we regressed the network created by neonates sharing cots against the network of putative TEs using exponential random graph models (ERGMs; Figure 4C). We constructed models in two stages, firstly to develop the overall model structure, only afterwards adding nodal covariates. Model B (edges + cots), was the best fit from the first stage (Supplementary Table 5), implying that cots influence transmission, and that the effect of time is explained by this effect of cots. When introducing nodal covariates, Model H (edges + cots + oxygen therapy + antibiotic therapy) was the final selected model and fit better than all other models tested using AIC, though had a slightly higher BIC than model F (which was the same, but without the oxygen therapy nodal covariate; BIC penalizes more for adding parameters), model diagnostics for Model H were within acceptable limits (Supplementary Figure 8). In this model a connection by cot increased the probability of a putative TE between two neonates from 0.00038 to 0.0036 (a 9-fold increase), one neonate in a pair receiving antibiotic therapy increased the probability of a TE to 0.0012 (a 3-fold increase) and one neonate in a pair being on oxygen increased the probability of a TE to 0.00062. Post-enrichment metagenomic data using these same models confirmed these effects (Supplementary Table 6), whilst sensitivity analysis using different SNP thresholds revealed that at low SNP thresholds (0 and 3) the number of putative TEs was too low to see a clear effect (Supplementary Table 6).

## Discussion

In this study we present multiple pieces of evidence that colonisation and infection of neonates by ESBL *Kpn* is related to within-hospital transmission and highlight the importance of IPC in the ward environment. Cots, ward surfaces, oxygen delivery equipment, sinks and mothers’ hands specifically are implicated, and it is likely that IPC practices that do not contain neonatal stool effectively act as the primary driver.

Previous studies at QECH have shown high hospital-associated ESBL colonisation in adults^21^. Here we show that neonatal colonisation with ESBL bacteria occurred rapidly, with *Kpn* the most frequent species identified despite exposure to multiple ESBL organisms from the ward environment, particularly *A. baumannii*, while mothers were more likely to be colonised with ESBL *E. coli* than *Kpn*. ESBL *Kpn* may be adept at colonising the ward environment, giving *Kpn* greater opportunity to be presented to the neonatal mouth and/or that neonates have poor colonisation resistance in the first few days of life due to an immature microbiome, that *Kpn* is able to exploit^22^.

Despite high numbers of *A. baumannii* isolated from ward surfaces, relatively few episodes of colonisation and invasive infection with *A. baumannii* are observed in our setting^16^ in contrast to sites in other countries such as South Africa^1^ or India^23^. This may be because highly invasive procedures (long lines, central lines, ventilation) that are risk factors for *A. baumannii* infection are not performed on Chatinkha NNU. Other studies have shown large variations in inpatient neonatal intestinal colonisation with *Acinetobacter baumannii* depending on country and reported resistance pattern, from 14-57%^24,25^.

Risk factors for neonatal colonisation were female sex, receipt of oxygen, caesarean delivery and antibiotic administration. Re-usable oxygen delivery equipment represents a potential point of entry for ESBL organisms and coupled with the results of the ERGM model and the transmission analysis (using the post-enrichment metagenomic data) indicates this may be an important mode of transmission of ESBL-*Kpn*. Ensuring that these items are appropriately cleaned or not re-used is vital. Both female sex and antibiotic administration as risk factors for neonatal colonisation with ESBL-*Kpn* have been shown before^26^. Antibiotics likely perturb the neonatal microbiome, favouring colonisation by ESBL-*Kpn*. Caesarean delivery is known to affect the microbiome and increase *Kpn* colonisation^27^.

Temporal analysis of the relationship between STs circulating on the ward and invasive disease indicated potential synergy; STs responsible for invasive disease and stool colonisation were co-circulating in the ward environment immediately before and directly following invasive and colonisation cases. This might represent a positive feedback cycle in which *Kpn* is excreted in large numbers by colonised neonates and may explain why outbreaks of *Kpn* neonatal sepsis occur on neonatal wards, including at this site^5, 28^. The source attribution model and analysis of bacterial circulation around the ward indicated the greatest relationship between neonates’ stool and cots, mothers’ hands and swaddling cloths, all areas that are easily contaminated by neonates’ stool.

Combining genomic and temporal data uncovered transmission to neonates primarily from cots, ward surfaces, and other neonates. Post-enrichment metagenomics particularly highlighted the role of ward surfaces, including sinks and oxygen delivery equipment. ERGMs confirmed the role of cots in transmission, as well as the role of oxygen delivery equipment and neonatal antibiotics. Neonate stool and compartments in close contact with the stool were important in transmission. Due to resource constraints, neonates are not swaddled and do not have nappies but lie on swaddling cloths, and mothers are responsible for laundering these items. Stool management interventions, which may involve use of disposable diapers or hospital linen services, should be further investigated^29^. This observation may partly explain the effect of Kangaroo mother care (KMC) in reducing neonatal deaths from infection, as neonatal stool is well contained^30^.

A limitation of this study is the small number of participants, and the low proportion of neonates on the ward at any one time recruited into the study. The low numbers reduced our power to detect small effects, and the low recruitment density (there are ~5,000 admissions per year to the ward and we recruited 94 neonates over eight months) would have decreased our ability to detect transmission events between these neonates. The number of connections between nodes in a fully connected network is represented by the equation n(n - 1)/2 and the number of connections thus increases in a quadratic manner as the nodes increase. Hospital transmission studies can be biased by the timing and location of sample collection, a fact that we tried to ameliorate by sampling many locations and performing multiple different analyses to confirm our findings.

We highlight that ESBL-*Kpn* in Malawi are primarily transmitted to neonates from the ward environment, including cots, ward surfaces, oxygen delivery equipment, sinks and maternal hands. Neonates themselves may be acting as amplifiers by excreting *Kpn* in their stool. These findings highlight IPC interventions that should be explored to prevent transmission of ESBL-*Kpn* in similar contexts. These include use of single-use oxygen tubing, effective ward surface decontamination, reductions in cot-sharing, replacing hard to clean wooden cots with wipe-clean ones, maternal hand hygiene interventions and interventions to improve handling of neonatal stool such as nappies or linen services.

## Data Availability

All data produced in this manuscript will be available at https://gitlab.com/ohapearse/cres and https://github.com/ohapearse/neonatal-kleb-colonisation-transmission.

## Acknowledgements

First, we would like to thank the mothers and babies who were involved in the study and the clinical team that looked after them. We would like to acknowledge the work of the MLW microbiology laboratory team, who identified and stored the invasive isolates. We would like to acknowledge Barry Rowlingson for preparing the QR codes that were used to track the movement of the neonates. We would also like to acknowledge Core Sequencing and Pathogen Informatics teams at the Wellcome Sanger Institute for their support with the bioinformatics.

## Author contributions

The study was conceived by RL, NAF, JC, and CJ. Methodology was developed by NAF, CJ, RL, PM, TE, EH, PS, JC and OP. Software was developed by OP and CJ. Validation was performed by OP and AZ. Formal analysis was performed by OP, EH and CJ. Investigation was performed by HM, OP, PS and AZ. Resources were provided by NAF, NRT and CJ. Data curation was performed by OP. Writing the original draft was performed as part of OP’s PhD by OP, NAF, CJ, JC and EH. Reviewing and editing the manuscript was performed by all authors. Visualisation was performed by OP and EH. Supervision was performed by NAF, KK, ET, CJ, JC, EH and NRT. Project administration was performed by OP, RL and HM. Funding acquisition was performed by NAF.

## Supplementary Materials

### Supplementary Figures

**Supplementary Figure 1.**
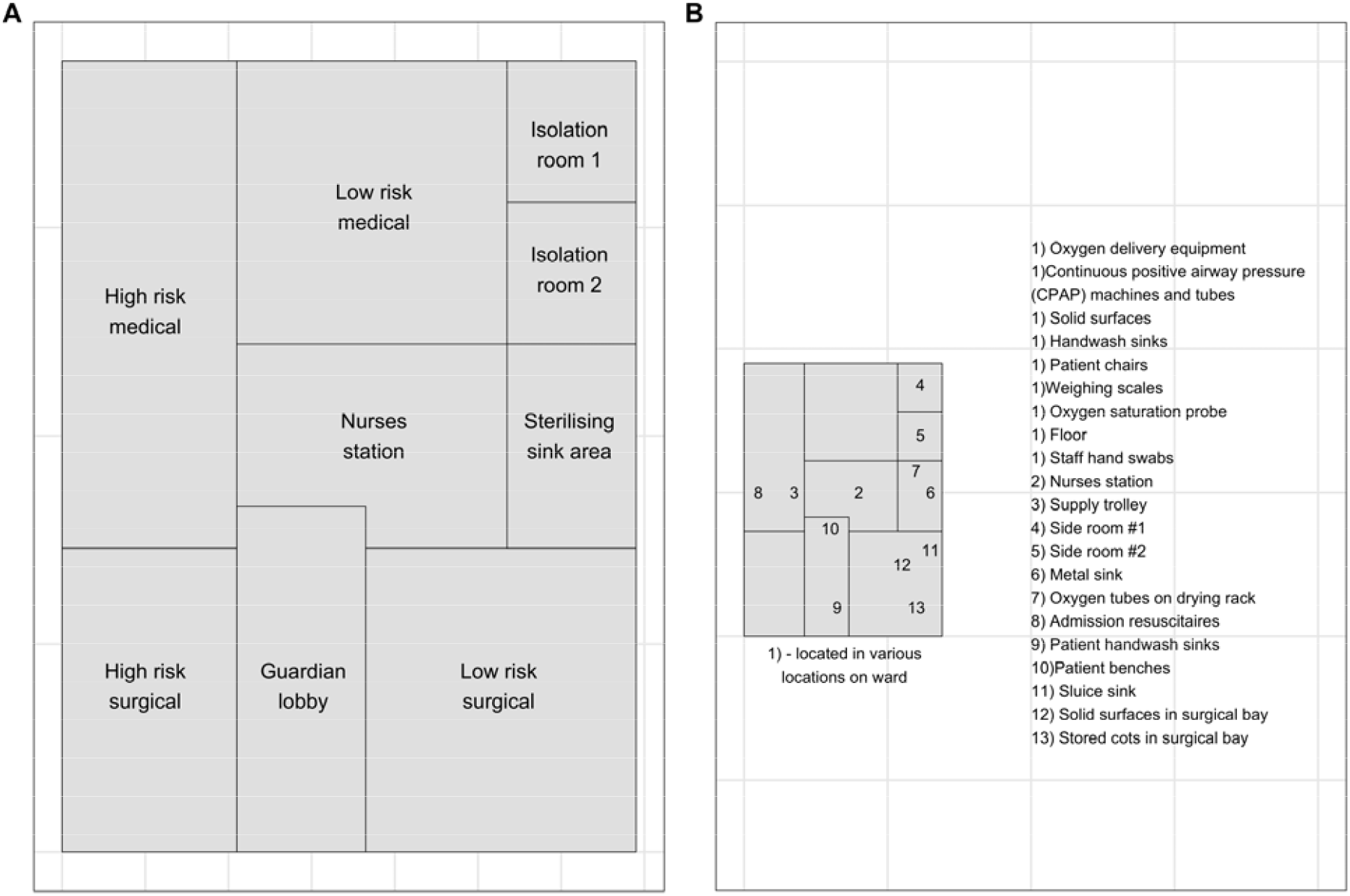
Two floor plans of the Chatinkha nursery. A) A floor plan showing the different areas of the Chatinkha nursery. B) A floor plan showing the locations of the different surface swabs, swabs that had multiple locations are marked as location 1). Other swabs are marked on the map with their approximate location.

**Supplementary Figure 2.**
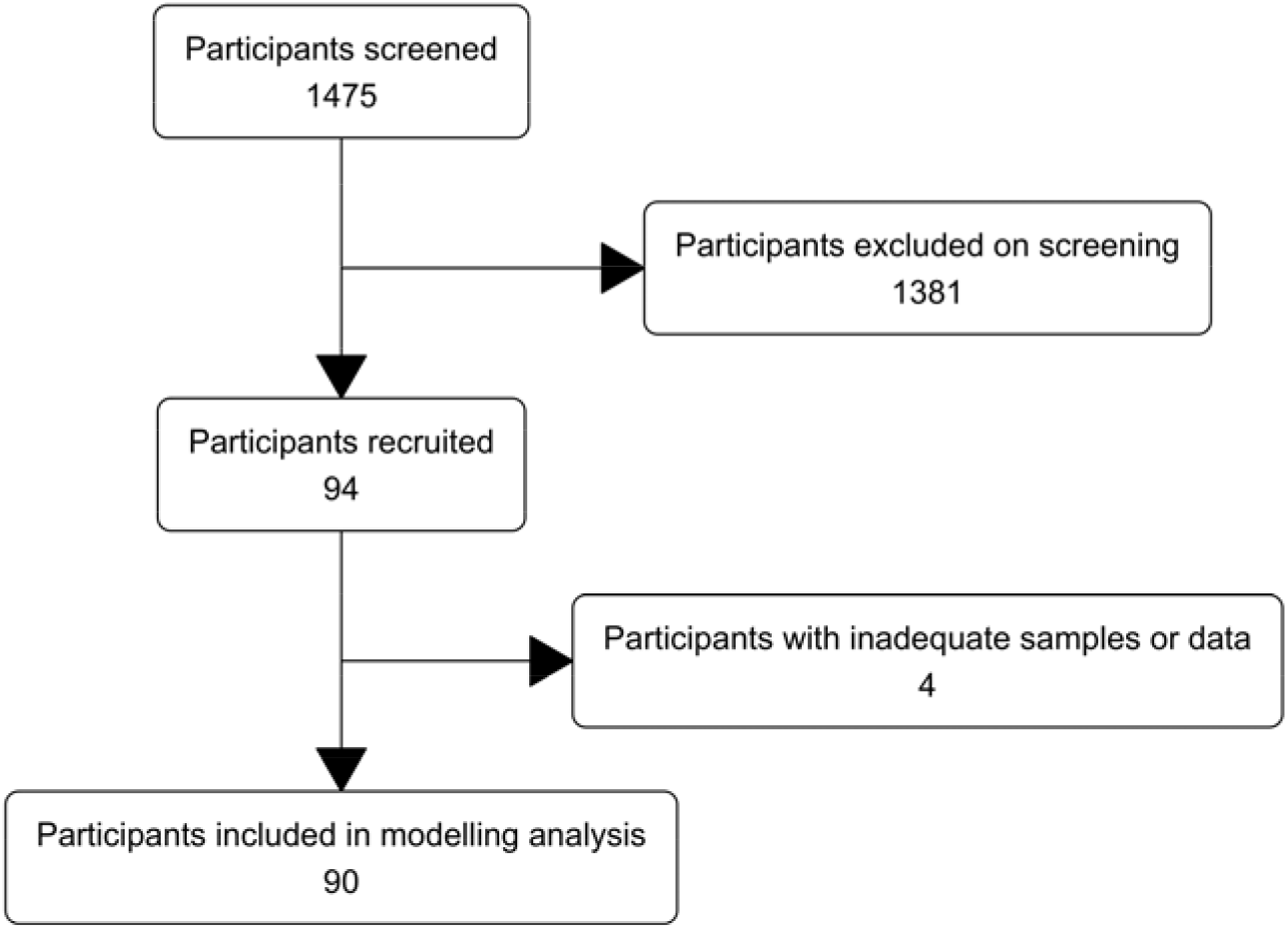
A flow chart showing the inclusion and exclusion of participants in the study.

**Supplementary Figure 3.**
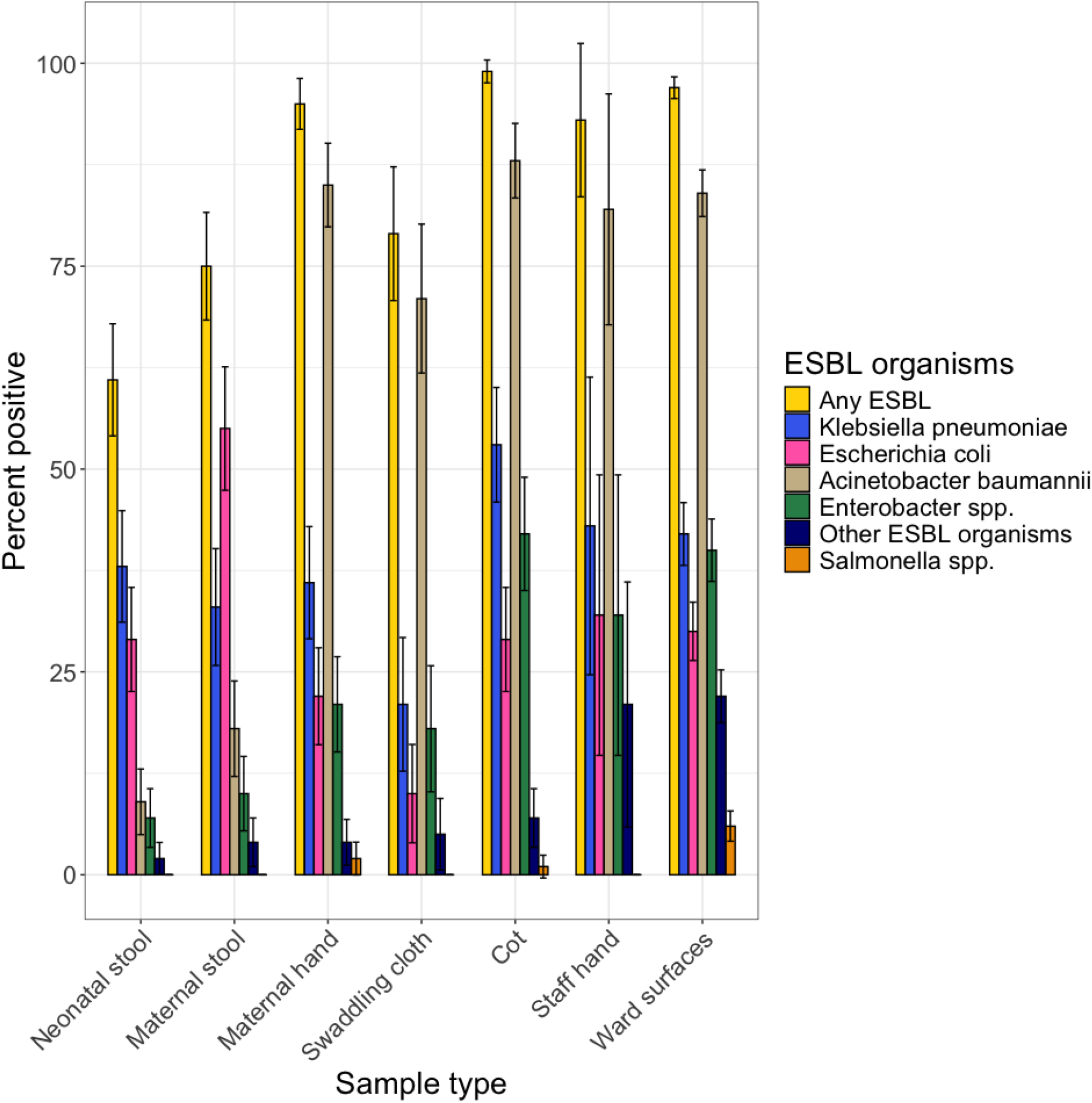
The percent of all samples that were positive for any ESBL organisms (yellow) or other specific organisms (other colours), by sample type.

**Supplementary Figure 4.**
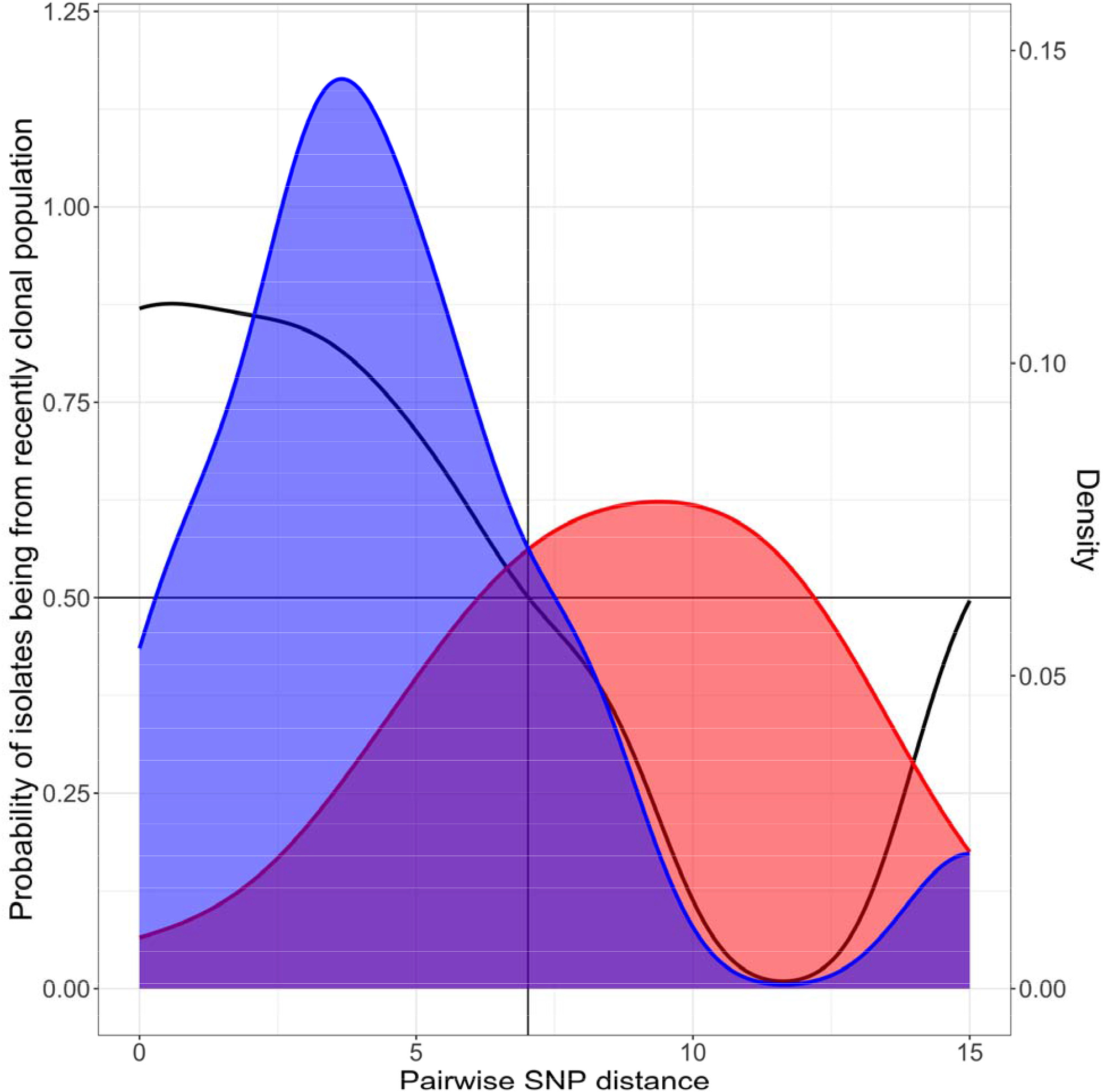
Density plots showing the densities of the pairwise SNP distance of isolates that are closely related (blue) and those that are not closely related (red). The intersecting straight lines represent the value at which an isolate is as likely to be recently clonally related as not. The black line represents the probability of two isolates being recently clonally related.

**Supplementary Figure 5.**
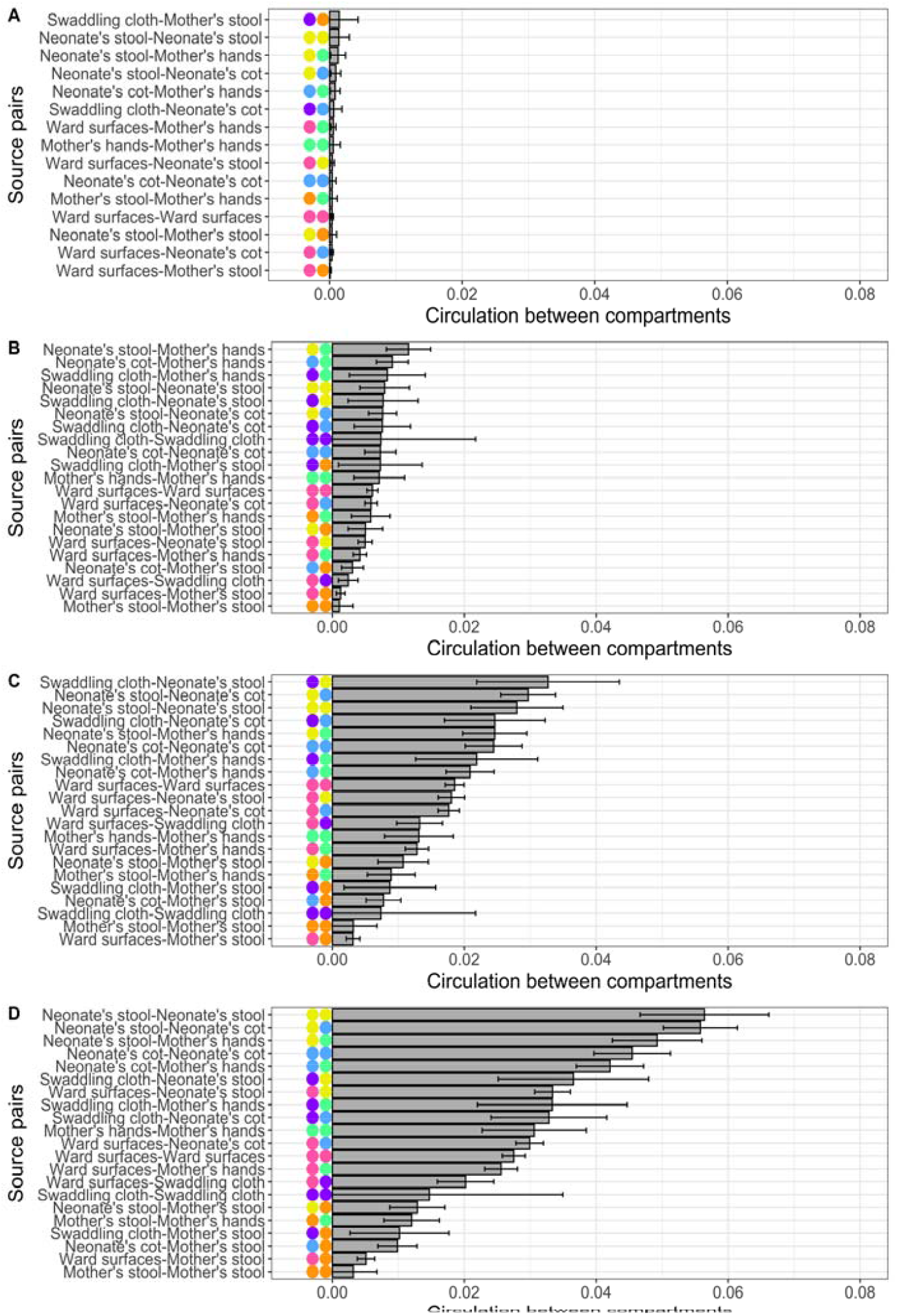
Circulation between the different ward compartments, as defined by different pairwise SNP distances for single colony WGS. Circulation is determined as the number of isolates between two compartments with a pairwise SNP distance below the threshold divided by the total number of potential connections between the two compartments. The colour of the points represents the two sources in the pair for that bar. A) A pairwise SNP distance of 0. B) A pairwise SNP distance of 3. C) A pairwise SNP distance of 7. D) A pairwise SNP distance of 20.

**Supplementary Figure 6.**
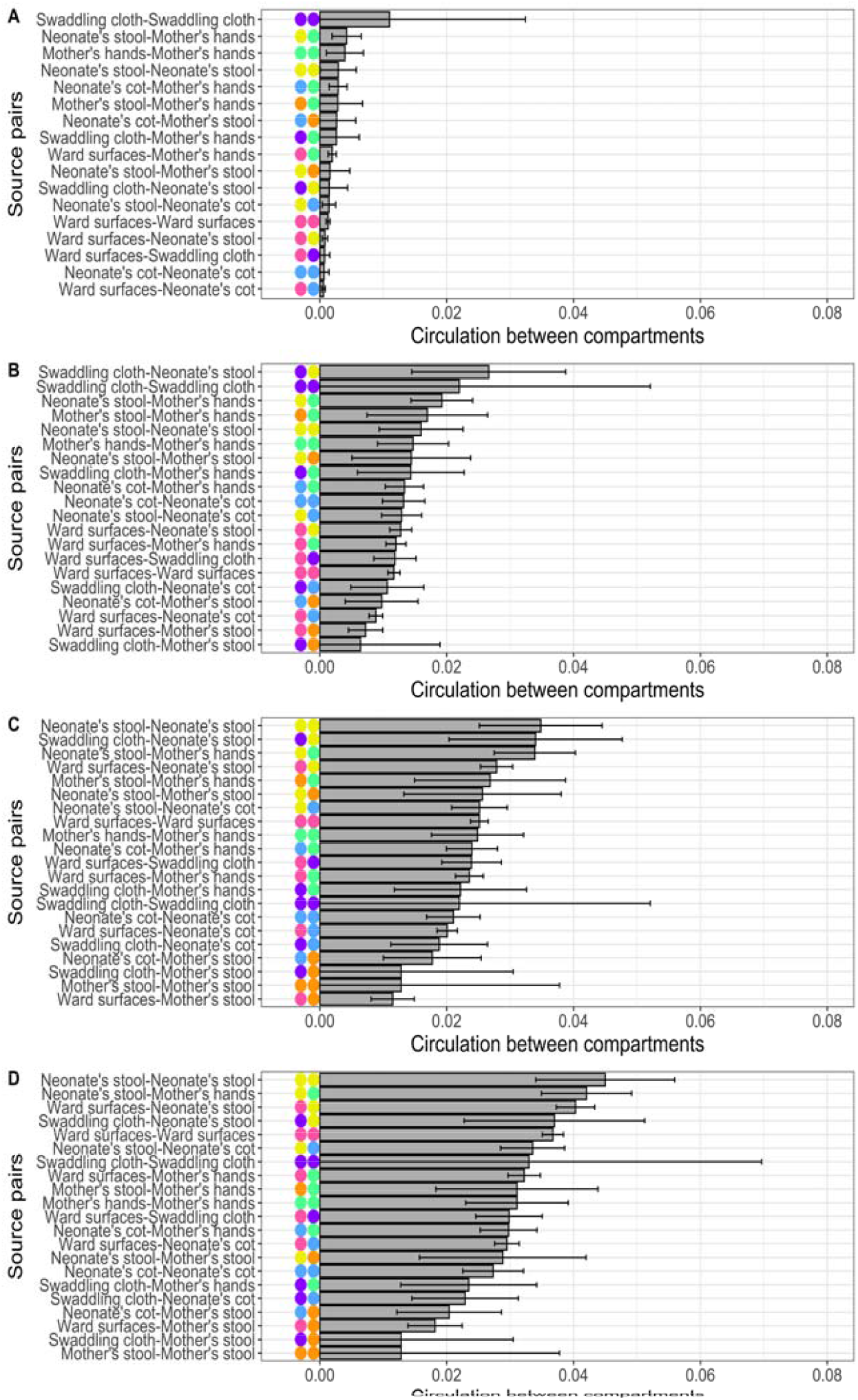
Circulation between the different ward compartments, as defined by different pairwise SNP distances for post-enrichment metagenomics (mGEMS). Circulation is determined as the number of isolates between two compartments with a pairwise SNP distance below the threshold divided by the total number of potential connections between the two compartments. The colour of the points represents the two sources in the pair for that bar. A) A pairwise SNP distance of 0. B) A pairwise SNP distance of 3. C) A pairwise SNP distance of 7. D) A pairwise SNP distance of 20.

**Supplementary Figure 7.**
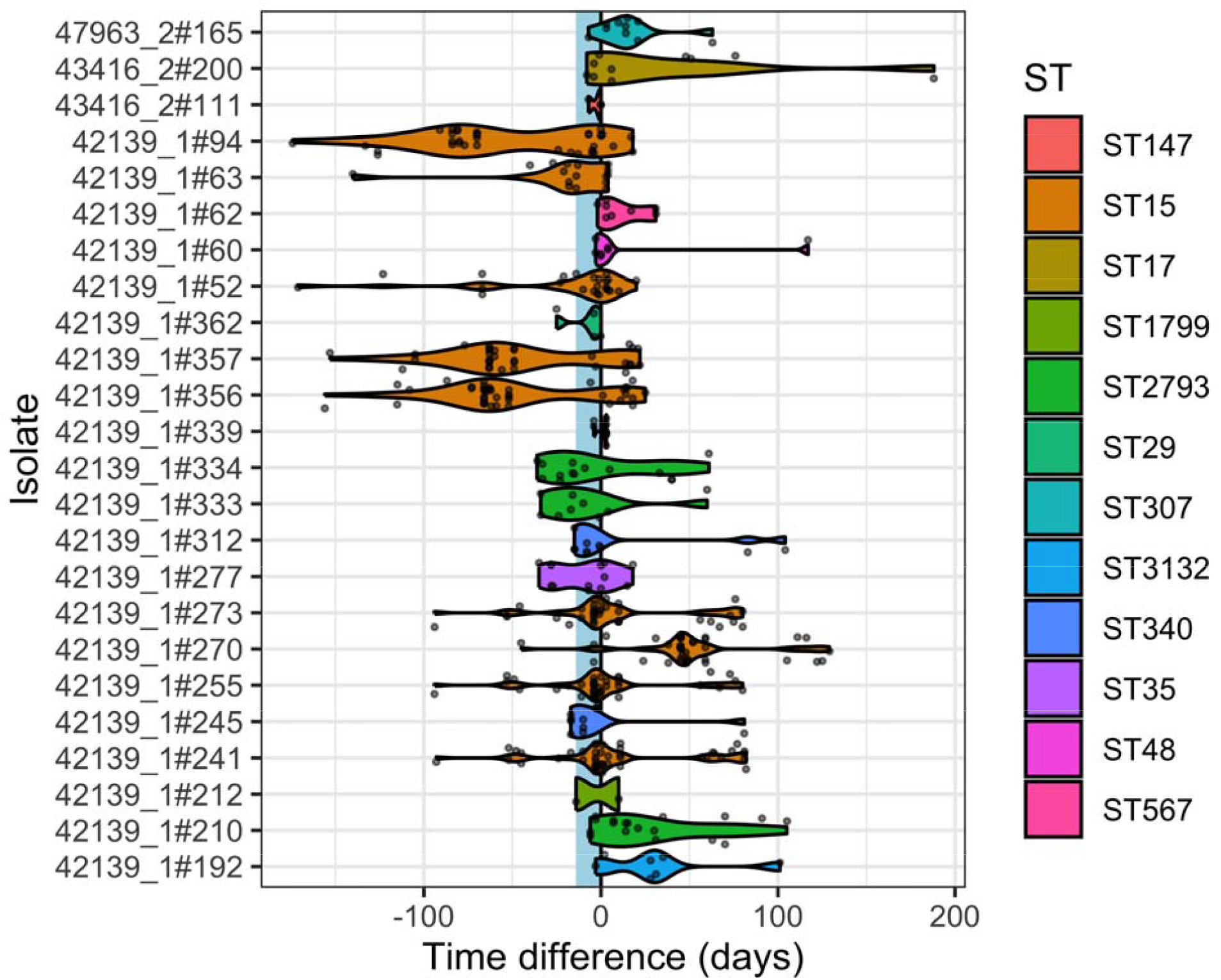
Violin plots showing the temporal occurrence of isolates on the ward in relation to isolates causing neonatal colonisation episodes (CEs). Isolates which had a pairwise SNP distance less than or equal to seven to the isolate causing the CE are plotted on the timeline in relation to the timing of the isolate colonizing the neonate, along with violin plots to represent the distribution of isolates in time. The different STs are represented by different colours. The light blue strip represents the time window in which an isolate had to appear to be classed as putative transmission.

**Supplementary Figure 8.**
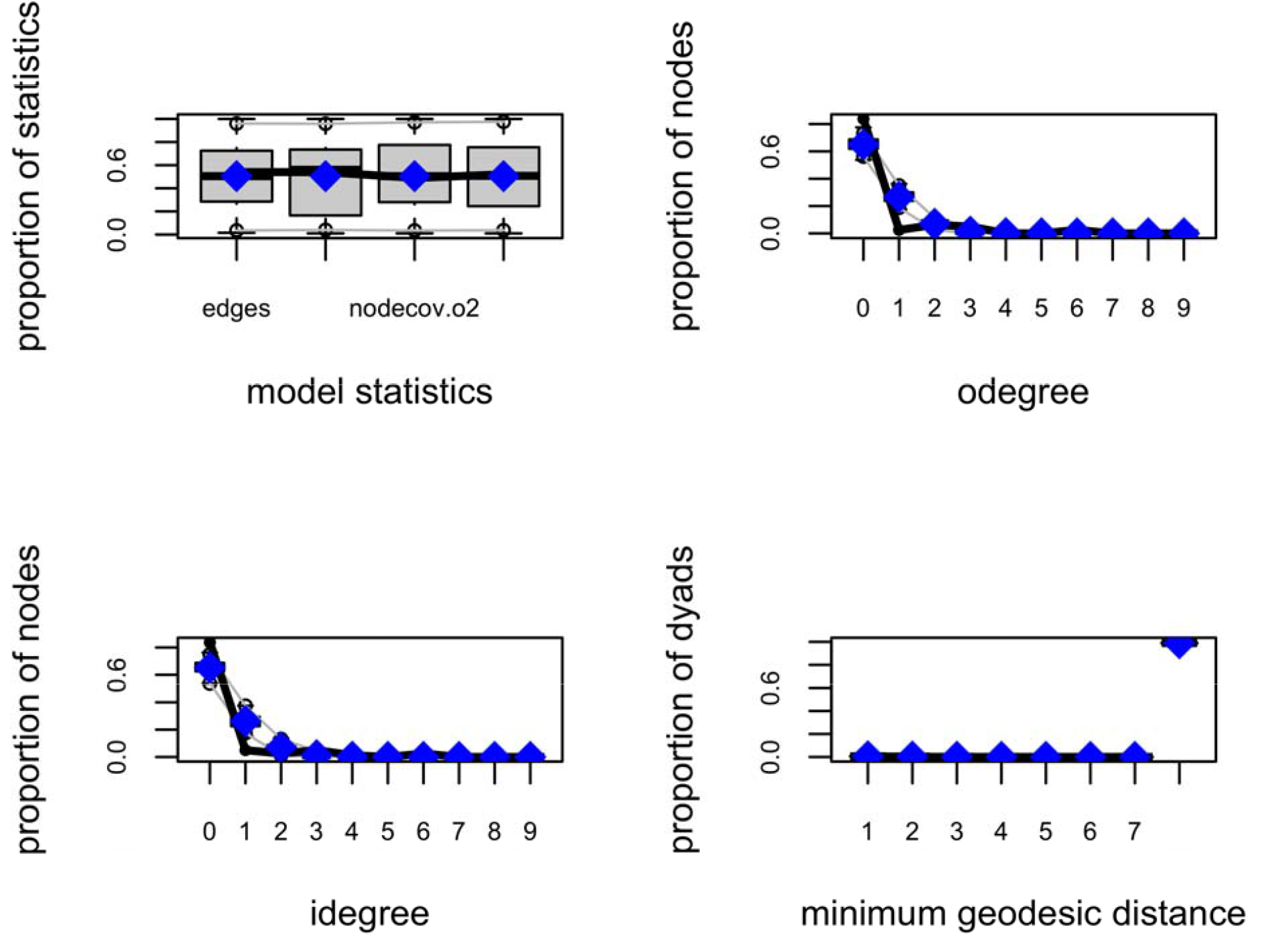
Comparisons of different model diagnostic metrics using simulations utilising the coefficients of the estimated Model H against the observed model statistics in the data. A) Model statistics, B) Outdegree (number of nodes with arrows directed away from them). C) Indegree (number of nodes with arrows directed towards them) and D) Minimum geodesic distance.

**Supplementary Figure 9.**
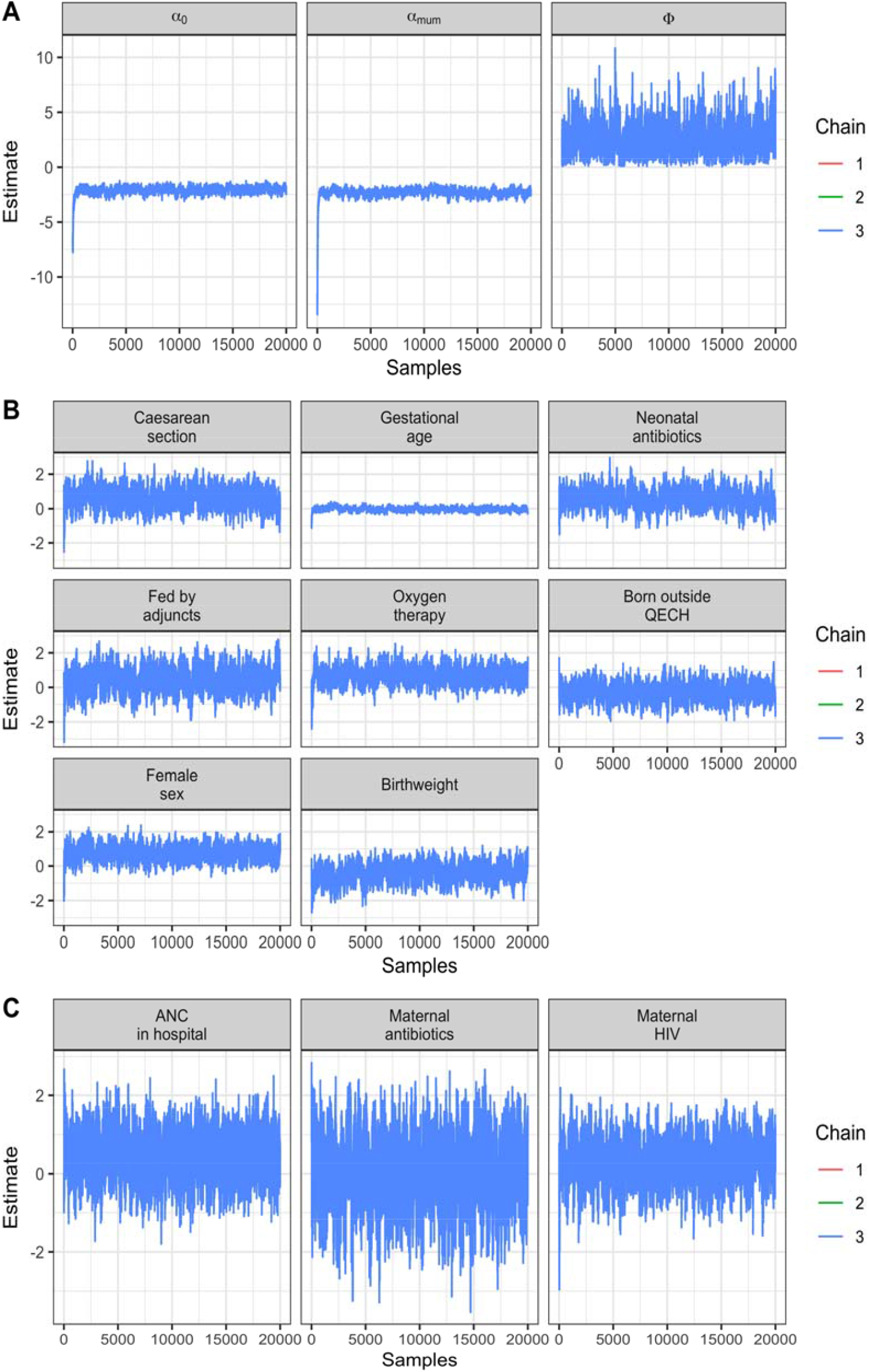
Trace plots of MCMC chains for the state transition model. A) For generic parameters, B) For neonatal covariates, C) For maternal covariates.

**Supplementary Figure 10.**
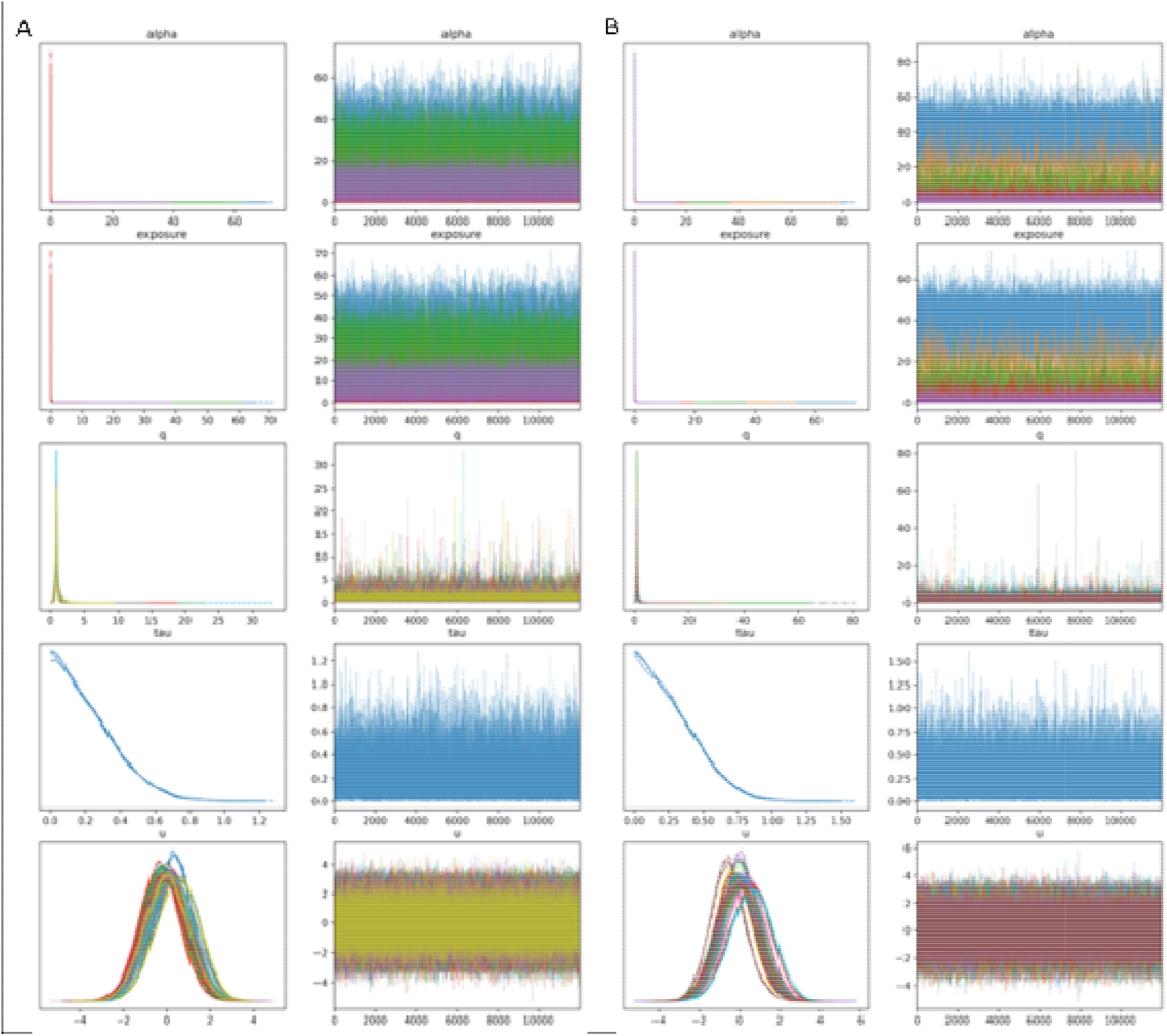
Trace plots of MCMC chains for the source attribution model. A) Single colony WGS data B) post-enrichment metagenomic data.

### Supplementary methods

In this section are detailed the specifics of the methodology used in this study.

#### Sample collection procedures

Stool samples were scooped into clear sterile 30ml plastic stool pots (Sterilin, UK). If stool sampling was not possible, then a sterile rayon-tipped swab (Medical Wire, UK) was inserted into the rectum, rotated for 10 seconds and stored in Amies gel media for transport to the laboratory.

Ward surface swabs, hand swabs and cot swabs were taken using a sponge-stick in 10ml of neutralising buffer (3M, UK). Oxygen tubing was swabbed using a small rayon swab in neutralising buffer. Using aseptic non-touch technique the swab was wiped over the relevant surface. We aimed to cover as much surface area as possible, so for small items of equipment we swabbed the whole surface. For medium items of equipment we swabbed all the high contact surface. For large areas such as floors we swabbed a random and representative area of the entire surface. For staff members, each week a single pooled hand swab was taken from at least five staff members. For the oxygen tubes good coverage of the inside surface of the tubes was attempted.

#### Study isolate identification and processing

Ward surface, cot, hand, and swaddling cloth swabs were incubated in buffered peptone water (BPW) for 24 hours; stool samples and rectal swabs were processed directly, as described^2^. All samples were plated on MacConkey agar and ESBL-selective chromogenic agar (CHROMagar, USA). Study and clinical isolates were identified and processed following protocols in the supplementary methods.

Pink colonies on chromogenic agar were assumed to be *Escherichia coli*, white colonies were assumed to be *Acinetobacter baumannii* unless they had the appearance of *Salmonella* spp., in which case they were identified using PCR. Blue colonies underwent PCR to determine whether they were *Klebsiella pneumoniae, Enterobacter* spp. or another member of the Group V Enterobacteriaceae. All identified isolates were stored, as were plate sweeps of the MacConkey plate.

For PCR a single colony from each relevant isolate was taken and DNA 95°C in molecular grade water for five minutes. *Kpn* and *Enterobacter* spp. primers were multiplexed. Samples underwent PCR as previously described^2^. The primers used for *Kpn* are based on those previously published^1^ (Supplementary Table 7).

All *Kpn* isolates were selected for sequencing. They were purity plated on MacConkey agar twice before 20-hour incubation in BPW. Dyad-associated samples and most ESBL-positive *Kpn* or *E. coli* ward surface swabs also had stored MacConkey plate sweeps sent for sequencing after 4-hour BPW incubation. BPW from all samples was centrifuged at 3500 rpm for 20 minutes to create cell pellets for DNA extraction.

#### Clinical isolate identification and processing

Routine, quality-assured diagnostic blood culture services have been provided to neonates on the unit by MLW since 1998 (ISO15189 accredited since 2019). For neonates with clinical signs of sepsis or meningitis a blood culture and CSF sample was collected. For blood cultures, 1 – 2ml of blood was collected using aseptic methods and inoculated into a single aerobic bottle (BacT/Alert, bioMérieux, Marcy-L’Etoile, France). These were incubated using the automated BacT/Alert system (bioMérieux, France). CSF samples were processed for cytology and biochemistry before also being incubated. Samples that flag positive were Gram stained and Gram-negative bacilli are identified by Analytical Profile Index (Biomérieux). Antimicrobial susceptibility testing was determined by the disc diffusion method (Oxoid, United Kingdom), following the current version of the EUCAST guidelines. All clinical *Kpn* isolates from Chatinkha NNU over the course of the study and 1 month afterwards were analysed as part of this study.

#### Sequencing and bioinformatics

DNA was extracted either manually using QIAamp DNA Mini kits (Qiagen, Germany) or the QIAsymphony DSP Virus/Pathogen kits on the QIAsymphony automated system with on-board lysis, following manufacturer protocols. Samples yielding <200 ng DNA were re-extracted. Adequate samples were shipped to WSI for sequencing.

Isolates were prepared using the Illumina protocol and sequenced on the Novaseq SP with 150 bp paired-end reads at 364-plex. Plate sweeps were similarly prepared and sequenced on the HiSeq 4000 with 150 bp paired-end reads at 96-plex.

#### Bioinformatic analysis

Full genomic bioinformatics analysis details are provided in the following paper^2^. Single colony WGS were assembled with SPAdes v3.10.0^3^, species identity was confirmed using Kraken v1.1.1^4^ and genome assembly statistics were checked. Any sample with greater than 5% read content other than *K. pneumoniae, K. variicola* or Unclassified was excluded, as well as samples with <20 or >150 contigs, genome size of <5MB or >6.5MB, or greater than 5% heterozygous SNPs. Sequence types (ST) were determined with Kleborate v3.0.0^5^. For single colony WGS isolates Snippy v4.6.0^6^ was used to align the reads to the *K. pneumoniae* reference genome (accession number CP000647.1)^7^. Gubbins v3.2.1^8^ was used to exclude the hypervariable regions (representing recombination and phages) of the genome. Snpdists^9^ was used to determine the pairwise SNP distances between pairs of isolates.

Following single colony whole genome sequencing and subsequent quality control, 37 isolates were excluded, ten had organisms other than *Kpn* in the sample, 30 due to failed assembly or poor assembly metrics, one due to within-species contamination and five invasive isolates excluded as they were ceftriaxone susceptible; with some isolates excluded for more than one reason, leaving 597 colonisation or environmental isolates and 17 invasive isolates (614 total; here^2^ for colonisation and environmental isolate details and Supplementary table 8 for invasive isolates). In the case of multiple colony phenotypes resulting in more than one pick per plate, duplicated STs were removed, leaving 552 isolates in total.

For the post-enrichment metagenomic data processing a custom database was created, containing 1835 *Kpn* genomes and 1966 genomes of other bacterial species. The database was based on the one used in the original mSWEEP publication^10^ and the full details of the database are described elsewhere^2^. The *Campylobacter jejuni* and *Staphylococcus epidermidis* genomes were removed, leaving the *K. pneumoniae* database and *E. coli* database plus individual examples of other bacterial species. To these we added 462 *K. pneumoniae* single colony WGS isolates analyzed in this study, as well as a collection of 429 *E. coli* genomes from QECH. The entire database included 1835 *K. pneumoniae*, 1940 *E. coli* and 26 other organisms. The labelling for the mSWEEP database included clonal complexes and STs, so all sample labels were converted to multi-locus sequence type.

772 plate sweeps from MacConkey agar were selected for post-enrichment metagenomic sequencing. All 772 metagenomic samples were used for diversity estimation in the samples (mSWEEP). Of these, 638 resulted in a prediction of at least one *Kpn* ST using mSWEEP. *Kpn* bins that were at greater than 10% relative abundance in the sample were selected for further processing. mGEMs v1.0.0^11^ was run on the raw post-enrichment metagenomic reads. Bins were assembled using Shovill v1.1.0^12^. Pseudo-assemblies with a total genome length not between 5 and 6.5MB were excluded, as were pseudo-assemblies with greater than 1000 contigs. Kleborate v3.0.0^5^ was then run on these pseudo-assemblies. Further QC was undertaken by checking for concordance between the predicted ST of mSWEEP and ST called by Kleborate, with pseudo-assemblies being excluded if there was discordance. Pseudo-assemblies were processed using the same pipeline for SNP distance determination as for the single colony WGS.

#### State transition model

We consider each neonate and mother to exist in one of two mutually exclusive states, either uncolonised (*U*) or colonised (*C*). We denote 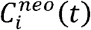 and 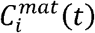 as a neonate and mother ‘s colonisation status respectively at time *t*, taking the value one if colonised and zero otherwise. For convenience, the collection of neonate and maternal colonisation statuses are represented below as the vectors 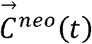 and 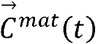.

To set the initial conditions for the model, we assumed that at birth (*t* = 0)a neonate is uncolonised, however at the time of birth mothers are assumed to be in the same state as their first observation. Each time step, or day, both neonates and mothers have a risk *P*_*uc*_ of transitioning from *U* → *C*. We are interested in estimating the rate of transition from *U* to *C* (*λ*), and the effect of patient level covariates in modulating this rate.

#### Transition rate and probability

We assume that neonate *i* transitions from *U* → *C* at rate *λ*^*neo*^(*t*) such that

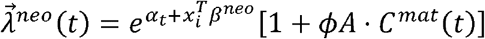

*α*_0_ is the baseline log hazard rate of colonisation. 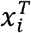 represents the neonatal covariates, with *β*^*neo*^ representing their coefficients. The coefficient *ϕ* represents the force of colonisation from mother to neonate, and *A* is an adjacency matrix connecting neonates to mothers such that *a*_*ij*_ = 1 if neonate *i* is a child of mother *j*, and *a*_*ij*_ *= 0* otherwise.

For mothers the transition rate 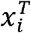 is defined as:

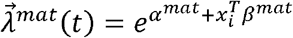

Where *α*_*mat*_ represents the baseline log hazard rate of maternal colonisation, represents the maternal covariates and *β* _*mat*_ represents their coefficients. Note that this implicitly assumes that mothers cannot be colonised by neonates.

To account for uncertainty in testing, we assume that our observed colonised status is *Bernoulli*-distributed conditional on the modelled colonisation status such that:

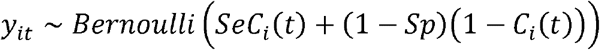

Where *y*_*it*_ is the observed colonisation state for individual *i* at time *t* (i.e. the test result),*Se* represents the sensitivity of our testing and *Sp* the specificity.

Parameter inference was carried out using a Bayesian approach, with data-augmentation Markov-chain Monte Carlo (MCMC) methods to marginalise over the unobserved neonate and maternal transition times^13^. The joint distribution of the parameters and unobserved data is estimated by drawing samples using a Metropolis-within-Gibbs MCMC algorithm^14^.

Priors were set as:

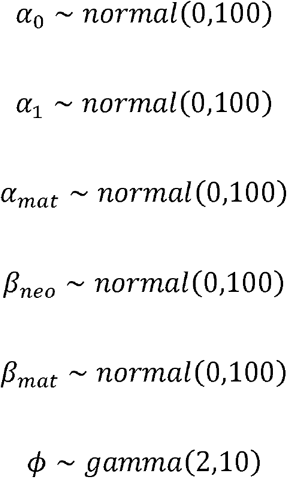

The MCMC algorithm was run on three independent chains starting from different values, for 20000 iterations, with the first 2000 discarded for burn-in. The convergence of the chains was assessed by inspecting the trace plots, to ensure that they were tracing the same distribution after burn-in, and by calculating the Gelman-Rubin statistic^15^ (Supplementary Figure 9; Supplementary Table 9).

This analysis is built on the gemlib package^16^, which is a Python library for data-augmentation MCMC on discrete-time epidemic models. The package is in development. The code is available on Gitlab^17^.

#### Covariate definitions for the risk factor model

- Fed by adjuncts – defined as fed by any other method than breastfeeding, i.e. orogastric tube or cup feeding;
- Oxygen therapy – administration of oxygen by any delivery device (oxygen tubing and CPAP tubing was re-used);
- Neonatal antibiotics – administration of antibiotics to the neonate during or before the study period;
- Intra-partum antibiotics – administration of antibiotics during labour;
- Pre-partum antibiotics – administration of antibiotics to the neonate’s mother within the four weeks before delivery;
- Maternal HIV – confirmed diagnosis of HIV in the mother;
- Born outside QECH – neonates born at a facility other than QECH.

#### Source attribution model

To quantify the contribution of different sample types to colonisation of neonates with *Kpn* we constructed a source attribution model. This is based on the modified Hald model^18^ and is similar to the approach used in the SourceR package^19^. This utilises case counts split by ST to determine the relationship between cases of ESBL *Kpn* STs in different sample types and in neonatal stool. This allows us to attribute the contribution of the different sample types towards neonatal colonisation, as well as whether there are specific STs which are more or less likely to colonise neonates.

The number of observed case counts of ST *i* are modelled as a Poisson random variable with mean value:

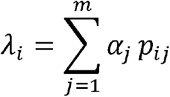

Where *p*_*ij*_ is the prevalence of ST *i* in source *j*, and *α*_*j*_ is the “source effect”, i.e. contribution to colonisation of source *j*. Sources described by *j* are; the general environment, neonatal cots, maternal stool, maternal hands, and swaddling cloths. Expressing ignorance as to which source was more important, priors for *α*_*j*_ for all *j* were set to:

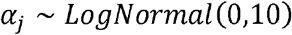

There is over-dispersion in the data, which may be because certain STs are better adapted to colonising neonates. To account for this, we also add a term for an STs propensity to colonise neonates *q*_*i*_ (or the “strain effect”). Thus, the equation becomes:

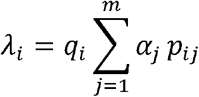

Having a term for every strain leads to over-specification of the model, as there are now *i* + *j* parameters, but only *i* observations. To deal with this, following one of the modifications to the Hald model, the size of *q*_*i*_ is restricted with the following expression:

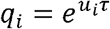

Where *u*_*i*_ is normally distributed with a mean of zero, a standard deviation of one and *τ* is a constant. This allows for the fact that most parameters may be equal but without forcing them to be so and replaces the need to identify parameters for strain effects with the need to identify a single parameter.

Here:

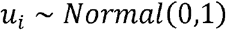

and *τ* is a constant with a prior distribution of:

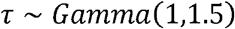

This approach shrinks the values of *q*_*i*_ to be close to one, reducing the effective number of parameters and aiding statistical identifiability.

The relative contributions of different sources *ζ* _*j*_ are calculated as:

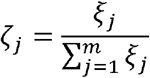

where ξ_*j*_ *is* the expected total number of cases from source *j*, i.e.

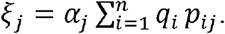

The model was run using PyMC^20^, with two chains run for 12000 samples and a burn-in of 1000 samples. Model fit was determined by inspecting trace plots (Supplementary Figure 10; all Gelman-Rubin statistics were < 1.01; Supplementary Table 10).

#### Genetic transmission analysis

To determine a SNP threshold consistent with transmission, pairs of isolates of the same ST colonising the same patient (assumed to be recently clonal) were compared to pairs of isolates from the same ST that were not colonising the same patient and isolated more than one month apart (assumed to not be recently clonal). Kernel densities of these two distributions were generated for SNP distances 0 - 15 and the probability of a specific SNP distance representing either a closely related population or not were calculated as *D*_*clonal*_/(*D*_*notclonal*_ + *D*_*clonal*_) where *D* represents the height of the kernel density. The SNP distance at which the probability of both isolates being from a recently clonal population was >0.5 was our defined SNP threshold (Supplementary Figure 4). Below this threshold the probability of two isolates being closely related was greater than the probability that they are not closely related. The SNP threshold that met this criterion was seven. All SNP analyses were repeated as a sensitivity analysis with SNP thresholds of zero, three and 20 (Supplementary Figures 5 & 6; Supplementary Tables 4 & 6).

Firstly, we looked at circulation of *Kpn* around the ward, charting the relationship between different ward compartments (neonatal stool, neonate’s cots, maternal stool, maternal hands, swaddling cloths and surface swabs). In the absence of evidence for specific transmission events it is useful to observe the level to which two sample types share bacteria as a proxy for circulation of bacteria on the ward.

The density of transmission between the two compartments was calculated as:

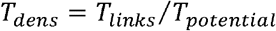

Where *T*_*dens*_ is the transmission density, *T*_*links*_ is the number of transmission links between two compartments and *T*_*potential*_ is the total number of potential transmission links.

When the transmission is within a single compartment (i.e.. Neonate’s stool to Neonate’s stool), *T*_*potential*_ is calculated as:

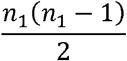

where *n*_1_ is the number of *Kpn* isolates in that compartment.

However, if transmission is between two different compartments (i.e.. Neonate’s stool to Neonate’s cot), *T*_*potential*_ is calculated as:

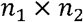

where *n*_1_ is the number of *Kpn* isolates in one compartment and *n*_2_ is the number of *Kpn* isolates in the other compartment.

The above approach does not give information about specific transmission events. To examine this, we looked at neonatal colonisation events (CEs) and utilised a time signal. Every neonate that was colonised with a *Kpn* of a specific ST was noted to have a CE. Neonates could have multiple CEs if they had been colonised with different STs. Each CE was treated separately. We looked at all isolates from samples taken between one and 14 days before the onset of each CE to see if there were any isolates which had a pairwise SNP distance lower than the SNP threshold. If they did, we classed this as a putative transmission event. We used this definition because finding a closely related organism in a ward area immediately before finding a related organism in a neonate, may imply directionality. We allowed multiple putative transmission events per CE. We performed sensitivity analysis with different SNP thresholds from zero to 20 (Supplementary Table 4).

#### Exponential random graph models (ERGMs)

ERGMs were created using the ERGM package^21, 22^ on networks created with the network^23^ package from the Statnet^24^ suite. We firstly defined a network of neonates that might plausibly be linked by transmission (*Edges*_*SNP*_), based on having a pairwise SNP distance between their colonising *Kpn* isolates less than or equal to the threshold defined in this paper (seven). The presence or absence of an edge between two neonates was regressed onto the following variables within an ERGM framework utilising the ERGM package:

- *Edges*_*prob*_ - represents the overall density of connections between neonates;
- *Edges*_*cot*_ represents a network of neonates connected by their contemporaneous or sequential occupancy of the same cot;
- *T*_*diff*_ is the difference in date of admission between the neonates;
- *Nodes*_*k*_ represent various nodal covariates.

Different combinations of these covariates were used, and model selection was undertaken to decide on the final model. The overall model structure was decided upon, using the fit metrics Akaike Information Criterion (AIC) and Bayesian Information Criterion (BIC). Firstly, we investigated the terms above without any nodal covariates.

Once the best fitting model structure was selected, important nodal covariates were added to see their effect on model fit. These were covariates that would plausibly influence the cots that neonates occupy (and may represent confounders) and/or the probability of becoming colonised with ESBL *Kpn*. These were oxygen therapy, neonatal antibiotics, and birthweight.

Coefficients from these ERGMs represent the change in the log-odds of a tie for a unit change in the predictor. The change in the log-odds of a tie can be transformed to a change in the probability of a tie for a unit change in the predictor by calculating the reverse log-odds of the coefficient.

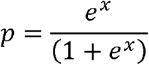

Where *p* represents the change in probability of a tie and x represents the change in log-odds of a tie. For the covariate cots, this was the change in probability when a pair of neonates were connected via cot, whilst for the nodal covariates this was presented as the change in probability if one neonate was exposed to the covariate.

